# Hormonal contraceptive drug use trends in the Estonian Biobank

**DOI:** 10.1101/2025.08.15.25333767

**Authors:** Jelisaveta Džigurski, Märt Möls, Kristi Läll, Hannah Currant, Mall Eltermaa, Estonian Biobank Research Team, Reedik Mägi, Lili Milani, Triin Laisk

## Abstract

**Importance:** Hormonal contraceptives (HCs) support reproductive goals and alleviate symptoms of gynaecological conditions for millions of women. Despite widespread use, individual long-term HC use trends, medication switching, and impact of genetics on HC side effects remain uncharacterised.

**Objective:** To study HC use trends among Estonian Biobank (EstBB) participants, compare with national statistics, and evaluate suitability of EstBB for studying genetic risk for HC side effects.

**Design:** Longitudinal descriptive analysis of HC refill data collected from 2004–2022 and user profiles, integrating demographic, genetic, and electronic health records data.

**Setting:** Volunteer-based longitudinal cohort representing 20% of Estonian adult population, where participants signed broad informed consent.

**Participants:** Women aged 15–55 in 2004–2022 with complete health and medication history available through linked electronic health records.

**Exposure:** HC use defined as prescription and purchase of drugs of the Anatomical Therapeutic Chemical (ATC) coding system level 3 G02B and G03A, and ATC level 5 G03HB01 during follow-up.

**Main Outcomes and Measures:** Primary measures included age-stratified annual HC users prevalence rates, inferred HC usage period duration, switching frequencies, and user profiles. Secondary measures included HC use during COVID-19 period, and genetic and health-relatedthromboembolism risk factors.

**Results:** Over 19-year study period, twenty HC formulations with five administration routes (intrauterine device, ring, pill, patch and implant) were used by 73,071 women (mean age at joining EstBB (sd) = 35.6 (10.6)). Comparable to the Estonian population, combined HCs dominated, while progestin-only HC use increased with age and time. Next, 64.3% of users switched formulations at least once, with 17.7% being rapid switchers. Rapid switchers showed side effect-related diagnoses before switching, indicating the dataset’s potential for studying genetic risk of side effects. Medical abortion overlapped with inferred usage periods in 3.2% of users, suggesting contraceptive failure. Finally, 5.3% of HC users carried at least one VTE-associated genetic variant, and 1.5% of carriers developed thromboembolism during inferred HC use.

**Conclusions and Relevance:** These HC use trajectories, consistent with population statistics, provide insights into real-world use patterns, offering additional context and support for understanding prescription trends, women’s preferences, as well as HC effectiveness and safety. Moreover, EstBB dataset has potential for genetic analyses of HC use and associated side effects.

**Graphical Abstract:** 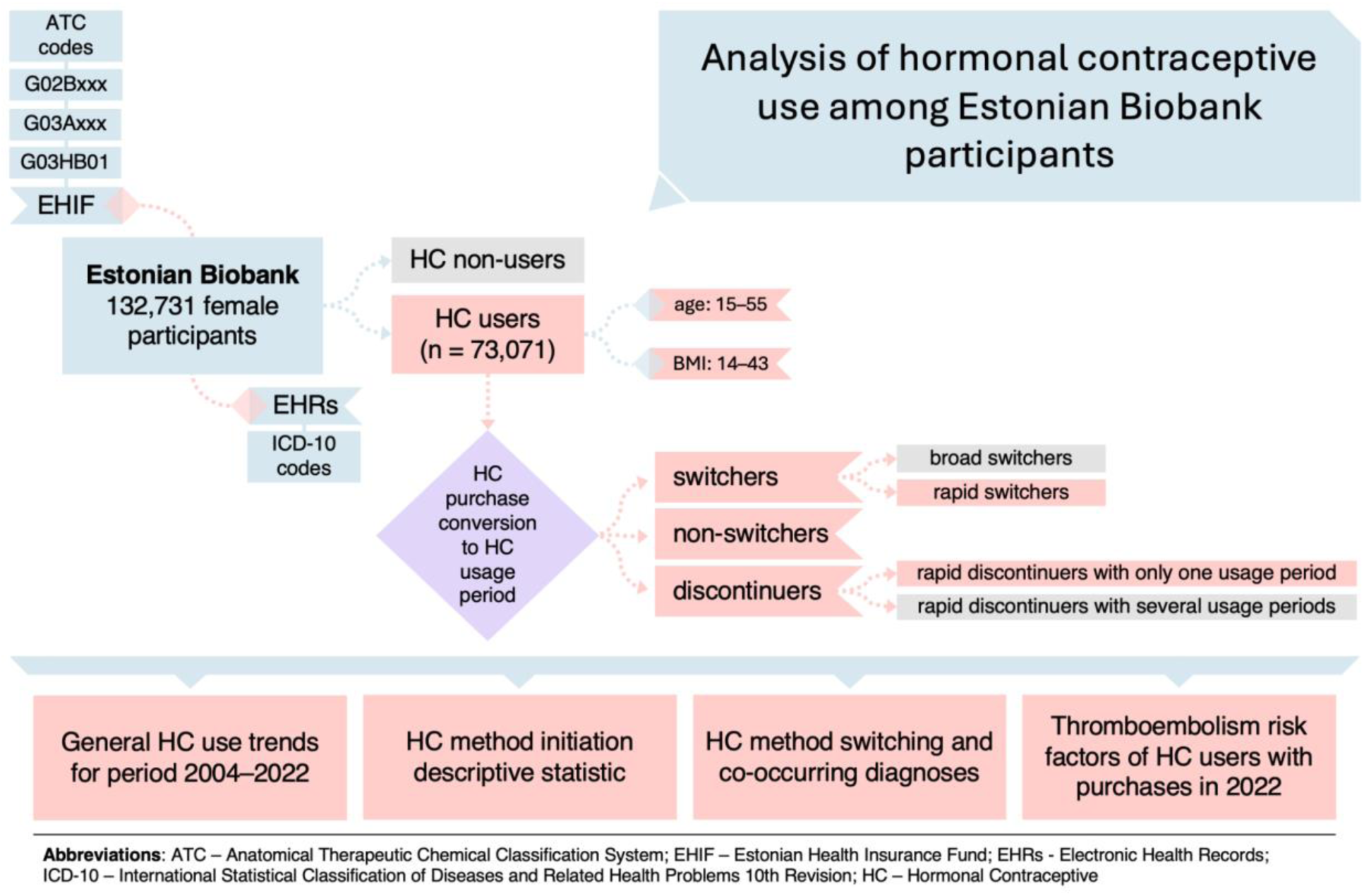

## Introduction

Hormonal contraceptives (HCs) are prescription drugs with approximately 300 million users worldwide^1,2^. HCs are mainly used to support women’s reproductive goals and treat symptoms associated with gynaecological conditions, such as polycystic ovary syndrome (PCOS)^3^, endometriosis^4^, and heavy menstrual bleeding^5^. Various HC formulations with different administration routes have been designed to meet women’s individual needs, overall health, and lifestyle. Generally, the active components are either a combination of progestin and estrogen (combined hormonal contraceptives, CHCs), available in fixed or sequential formulations, or progestin alone (progestin-only contraceptives, POCs). Each hormonal method is associated with different potential side effects and health risks^6^, which may lead to either discontinuation or switching of a medication^7–9^. Most notably, CHCs are associated with a slightly increased risk of venous thromboembolism (VTE) and arterial thromboembolism (ATE)^6,10,11^. Despite potential risks, the European Medicines Agency (EMA) has stated that since the risk of VTE with all CHCs is low, the benefits outweigh any potential risks; however, a woman’s risk for thromboembolism should be evaluated before prescribing CHCs^12^. Similarly, prescription recommendations exist for individuals experiencing certain conditions, such as migraine^13,14^. In parallel, continuous efforts are made to investigate and improve the safety and effectiveness of HCs^15–17^.

The availability of specific HC formulations and prescription patterns vary widely across countries, reflecting differences in healthcare systems and sociocultural norms^1,2^. The selection of the most suitable method is made according to the guidelines of the World Health Organization (WHO) and the Centers for Disease Control and Prevention (CDC), and based on the woman’s general health and clinical status^13,14^. A study published in 2021 found that during 2005–2019, the use of POCs increased in Estonia, while CHC use declined^18^. At the same time, it was found that there was a considerable number of women using CHCs who either had a history of thrombosis or multiple documented risk factors for thrombosis. Both VTE and ATE have several risk factors that, in theory, should be evaluated before prescribing HCs. These include family history, age and smoking status, but also underlying medical conditions, such as hypertension, cardiovascular disease, and others^19^. The use of certain drugs, such as antipsychotics or antidepressants, can also increase thrombosis risk^20,21^, although the results for antidepressants have been conflicting and could be due to side effects or drug-drug-interactions^22,23^. Moreover, VTE has well-established genetic risk factors, such as rare antithrombin, protein C and protein S deficiencies, and more common gain-of-function variants in the *F5* and *F2* genes (found in approximately 1–5% of individuals of European ancestry)^24–26^. The association of these common genetic variants with ATE events remains uncertain, with most studies showing modest or small effects^26–28^. Current WHO guidelines do not support routine screening for thrombophilia before prescribing HCs due to the relatively high test cost and low prevalence of these variants. However, a recent study using UK Biobank data suggested that evaluating the full spectrum of genetic risk for thrombosis, which is a polygenic condition, may improve risk stratification^29^. While this study highlighted the value of large population-based biobanks for the study of genetic risk factors associated with HCs, due to the age profile of the participants (40–60 years old at recruitment) and the nature of the study, the results apply mainly to the second-generation of oral HCs and relied on self-reported use, which does not provide comprehensive detail about prescription dynamics and users risk profiles.

The Estonian Biobank (EstBB) is a population-based biobank with ∼140,000 female participants, 43% currently aged under 50, for whom genotype and health-related data are available^30^. The electronic health records (EHRs) of EstBB participants are regularly updated by linking to the Estonian Health Insurance Fund (EHIF) and Estonian Health Information System, providing nearly complete coverage of diagnoses and prescriptions for all participants going back to the early 2000s when this type of information was digitalised. This rich data collection has formed the basis for studies exploring the genetic background of common traits, phenotypic comorbidities, and pharmacogenetics^31–36^. Given the large number of reproductive-aged women in the EstBB, and the detailed genetic, health, and prescription data available, EstBB is an ideal research setting for studying HC use and its pharmacogenetic aspects. However, previous studies have shown a “healthy participant bias” in population-based biobanks^37,38^, which may also affect contraceptive use and the presence of risk factors. Currently, we lack an overview of how the EstBB data compares to the Estonian general population regarding contraceptive use trends and risk factor profiles.

In this study, we characterise HC purchasing and usage trends among EstBB participants from 2004 to 2022. This unique cohort and longitudinal analysis provide a comprehensive understanding of prescription and purchase trends, as well as the individuals’ HC preferences, long-term use and switching dynamics. Additionally, we compare how the EstBB cohort reflects the observations on HC use from the general population, and evaluate the presence of thromboembolism risk factors among women using HC in EstBB to validate its robustness for pharmacogenetic studies.

## Materials and Methods

### Estonian Biobank

Estonian Biobank (EstBB) is a volunteer-based biobank with genotype, phenotype and biological data of more than 210,000 individuals who were 18 years and older at recruitment, with 65% being females^30^. The cohort represents 20% of the total adult population of Estonia. The ethical approval 1.1-12/624 for the study was granted by the Estonian Committee on Bioethics and Human Research (Estonian Ministry of Social Affairs), using data according to release application number 6-7/GI/16011 from the Estonian Biobank. All participants have signed a broad consent form. The comprehensive description of the cohort has been made in previous publications^30^. Briefly, the participants are volunteers who were recruited in 2002–2015 primarily by family physicians and in 2018–2019 via public events, media, and clinical lab visits. In addition to questionnaire data collected during the recruitment, health-related data are linked from the Estonian Health Information System, Estonian Health Insurance Fund, Cancer Registry, Tartu University Hospital Database, and many others. The information about individuals’ medical diagnoses and conditions dates back to 2004 and is stored according to the International Statistical Classification of Diseases and Related Health Problems 10th Revision (ICD-10 Version:2016) system. For the computational analyses carried out in this study, EstBB 206K data freeze - with follow-up until the end of 2022 - was used.

### Descriptive Data Analysis

#### Study participants

All female EstBB participants were included in the study, while the main focus was on those who had at least one record of an HC purchase between 2004 and 2022 and were aged 15–55 at the time of the purchase. Basic descriptive characteristics of HC users, such as age at contraceptive initiation, age at each HC usage period start date, and body mass index (BMI) closest to the usage period start date, were used for analysis. BMI values originated from self-reported data or have been extracted from EHRs, and were categorised by CDC criteria: (a) underweight (BMI < 18.5), (b) healthy weight (18.5 ≤ BMI < 25.0), (c) overweight (25.0 ≤ BMI < 30.0), and (d) obese (BMI ≥ 30.0).

#### Hormonal contraceptive usage

Data on HC prescriptions and purchases originates from the Estonian Health Insurance Fund, which subsidises medical expenses for people covered by national health insurance, and stores invoice information from treatments and drug purchases. The linking procedure has been previously described in Leitsalu et al^39^. Specifically, our dataset includes the purchase invoices from 2004 to 2022. For analysis, we used the Anatomical Therapeutic Chemical (ATC) Classification System code and the active substance name of the prescribed and purchased drug, purchased dosage, package contents, and purchase date.

HCs were defined as prescriptions of ATC level 3 G02B and G03A, and ATC level 5 G03HB01. There was no information about emergency contraceptives (ATC level 4 G03AD), as prescription is not needed for their purchase in Estonia. All entries where prescribed and purchased package ATC codes did not match were excluded from the analysis (n_purchases_ = 30,930 (1.8%)). In the following filtering step, all entries where the purchased dosage was less than 0.3 or greater than 6 were excluded (n_purchases_ = 653 (0.04%)). This ensured the dataset was free from mistype errors, as buying less than a third of a package is impossible. Gynaecologists prescribe per visit a maximum of three prescriptions, each for two drug packages (i.e. for a six-month coverage period). Finally, entries for which the individual’s purchase age was outside the 15–55 range (n_purchases_ = 1,306 (0.1%)) and individuals for whom BMI values closest to the HC usage start date were outside the range of 14–43 (n_users_ = 851 (1.1%)) were excluded.

The purchasing data was further converted into HC usage periods. Specifically, the purchased dosage and package contents for each HC method were used to calculate the total number of covered days per purchase, assuming that the contraceptive was used consistently and according to guidelines. The default number of covered days per purchase for pills, transdermal patches and vaginal rings was set to 28 or 35 (latter only for G03AC03), for implants to 1,095 days (corresponding to three years), while the coverage days for intrauterine devices (IUDs) depended on the formulation dosage (1,095, 1,835 or 2,190 days corresponding to 3, 5 or 6 years for 13.5mg, 19.5mg or 52mg levonorgestrel-releasing IUD, respectively). HC usage periods were additionally adjusted with ICD-10 pregnancy codes (O00–O99, *excl.* O85–O92 (complications predominantly related to the puerperium)) that occurred within the original coverage period. Regular purchases (<90 days of pause between two purchase dates) of the same HC were considered one usage period since the fecundity is generally restored in two to three months after discontinuation^40,41^. Furthermore, for HC formulations with intermittent dosing, the drug-free days were also considered as part of the usage period. Any purchase of a different HC formulation (different ATC level 5 code) would mark the start of a new usage period. There was no differentiation between brands, dosages, or regimes for the same HC formulation. The usage periods were created using Node.js v18.12.1^42^.

To answer different research questions, HCs were grouped into ATC levels 4 or 5 accordingly and categorised based on the hormonal composition, route of administration, duration of action and contraceptive method type. Combined oral contraceptives (COCs) were classified by pharmacological properties and progestin generations as: (a) anti-androgenic (cyproterone/ethinylestradiol, chlormadinone/ethinylestradiol), (b) second-generation (levonorgestrel/ethinylestradiol), (c) third-generation (desogestrel, gestodene, norgestimate, norelgestromin/ethinylestradiol), (d) fourth-generation (drospirenone, dienogest/ethinylestradiol), and (e) estradiol preparations (nomegestrol/estradiol, dienogest/estradiol valerate). While grouping COCs by generations is informal^43^, it is a common practice in the literature, where different COC generations are associated with different side effects profiles, especially thrombosis risk levels^44–47^. However, as it also varies across studies, we explicitly define our grouping criteria for clarity.

Finally, all individuals using HC were classified as switchers, rapid switchers, non-switchers and rapid discontinuers following these criteria: (a) switchers were individuals who used two or more different HC formulations during the study observation period, (b) rapid switchers were individuals who had short gaps (≤90 days) between two different formulation usage periods and short (<90 days) pre-switch formulation usage periods, (c) non-switchers were individuals who had long (>180 days) usage periods and used only one formulation during the study observation period, and (d) rapid discontinuers were individuals who had short (<90 days) usage periods and used only one formulation during the study observation period. Rapid discontinuers were additionally verified if they had any recorded diagnoses after the first purchase date, as evidence of not moving out of Estonia (which could otherwise result in missing prescriptions), and the purchase was made more than 90 days before the end of the study period (to rule out artefacts created by the end of the observation period). Overall, switchers are a wider class, with rapid switchers being its subset, while the remaining classes are mutually exclusive of each other and switchers (Supplementary Figure 1).

#### Longitudinal diagnosis history and thromboembolism risk factors

To identify clinical diagnoses before, during and after HC use, EHRs with ∼4,500 ICD-10 three-character category and four-character subcategory diagnosis codes were used. The illustration of the strategy can be seen in Supplementary Figure 2a. First, the records of clinical diagnoses in the three-month window preceding the first HC usage period start date were used to identify potential non-contraceptive reasons for HC initiation, such as D25 (Leiomyoma of uterus), E28 (ovarian dysfunction), E28.2 (polycystic ovarian syndrome), L70 (acne), N80 (endometriosis), N91 (absent, scanty and rare menstruation), and N92 (excessive, frequent and irregular menstruation).

Second, when evaluating diagnoses during the HC usage period and up to three months after the usage period end date to identify diagnoses related to potential side effects and possible reasons for HC discontinuation, we only considered first-time-ever diagnoses to avoid misinterpretation from pre-existing conditions. The same approach was used for diagnoses during the gaps between two different HC usage periods of rapid switchers. Our list of the curated selection of diagnoses related to potential side effects, based on side effects listed in patient information leaflets, included the following codes: B37 (candidiasis), F32 (depressive episode), F41 (other anxiety disorders), F52 (sexual dysfunction, not caused by organic disorder or disease), G43 (migraine), G44 (other headache syndromes), K80 (cholelithiasis), L70 (acne), L68.0 (hirsutism), N64.4 (mastodynia), N76 (other inflammation of vagina and vulva), N83 (disorders of ovary/fallopian tube), N84 (polyp of female genital tract), N92 (excessive, frequent and irregular menstruation), N93 (other abnormal uterine bleeding), N94 (pain and other conditions associated with female genital organs and menstrual cycle), R10 (abdominal and pelvic pain), R51 (headaches), and R53 (malaise and fatigue). Additionally, we investigated the occurrence of diagnosis O04 (medical abortion), which would indicate contraceptive failure.

Third, the prevalence of VTE/ATE was evaluated across the entire study period. VTE was defined using ICD-10 diagnosis codes I26 (pulmonary embolism), I80 (phlebitis and thrombophlebitis, *excl.* I80.0 (phlebitis and thrombophlebitis of superficial vessels of lower extremities)), I81 (portal vein thrombosis), I82 (other venous embolism and thrombosis), O22.3 (deep phlebothrombosis in pregnancy), O87.1 (deep phlebothrombosis in the puerperium) and O88.2 (obstetric blood-clot embolism), while for robust definition of ATE codes I21 (acute myocardial infarction), I24.0 (coronary thrombosis not resulting in myocardial infarction), I63 (cerebral infarction, *excl.* I63.6 (cerebral infarction due to cerebral venous thrombosis, nonpyogenic)), and I74 (arterial embolism and thrombosis) were used.

Expanding on the approach of Kurvits et al. (2021), in the subset of women who purchased HCs in 2022, thromboembolism risk factors were assessed in a two-year period ending on the first purchase date in 2022 (Supplementary Figure 2b). All thromboembolism risk factors were divided into three classes: (a) health conditions, (b) medication use, and (c) genetic risk factors. Health conditions were defined using the following codes: malignant neoplasms (C00–C97), sickle-cell disorders (D57), hemolytic-uraemic syndrome (D59.3), diabetes mellitus, type 1 and type 2 (E10–E14), obesity (E65– E66 or BMI ≥ 30.0), disorders of lipoprotein metabolism and other lipidaemias (E78.0–E78.5), migraine (G43), cardiovascular conditions (I05–I09, I20–I25 (*excl.* I21), I34–I39, I44–I50, I60–I70 (*excl.* I63)), hypertensive diseases (I10–I15), pneumonia (J12–J18), Crohn’s disease (K50), ulcerative colitis (K51), psoriasis (L40), rheumatoid arthritis (M05.8, M05.9, M06.0, M06.1, M06.8, M06.9), systemic lupus erythematosus (M32), and labour (O60, O80–O84) within 42 days before HC purchase^18,48^. Previous thrombotic events, family history of thrombotic events in first-degree relatives (parent–offspring or full siblings), and age >35 in 2022 were considered additional risk factors. The familial relationship between participants was inferred centrally for the EstBB. It is estimated based on coefficients calculated from the genetic data (kinship inference and identical-by-descent segment inference for close relatives) using the KING software, which implements a robust relationship inference algorithm^49^.

The concomitant use of other drugs associated with an increased risk of thromboembolism during the HC usage period was identified using purchasing data of ATC codes H02 (corticosteroids), N05A (antipsychotics), and N06A (antidepressants)^18^.

Additionally, the imputed genotype data were assessed for the presence of two known VTE risk-increasing variants (Factor V Leiden (FVL, rs6025, MAF = 0.02) in the *F5* gene and prothrombin G20210A (PTM, rs1799963, MAF = 0.01) in the *F2* gene). The genotyping and imputation procedure of the EstBB cohort has been described previously^30^. Briefly, all samples have been genotyped centrally using the Global Screening Array (Illumina) and imputed using a population-specific reference of 2,695 Whole Genome Sequencing samples^50^. Imputation INFO scores for both variants were >0.96, confirming the good quality of the genotypes.

#### Statistical analysis

The categorical data were summarised using frequency calculations, and the continuous data were summarised by calculating the mean and standard deviation. The annual use of HCs was calculated by summing up all active users within any given year. Annual HC users were stratified by five age groups (15–19, 20–29, 30–39, 40–49, and 50–55) using age at purchase calculated by subtracting the constructed birth date (YYYY-06-30, where YYYY is the year of birth, which is the only available data) from the purchase date. Constructed birth date was used to standardise age calculations across the study and minimise bias^51^. The age at the start/end of the HC usage period was calculated similarly to the age at purchase. The first-ever observed HC usage period during the study was considered the initiation period, and users were referred to as initiators. Annual prevalence rates were calculated by dividing the number of active users of the specific age group in the given year by the total number of women in the EstBB of the corresponding age group in that year. The data were analysed using *R base* and *tidyverse* packages in the R Statistical Software v4.1.3^52^.

## Results

### General use trends

The study cohort included a total of 132,731 women whose mean (sd) age at joining the EstBB was 38.3 (13.6) years. We focused on 73,071 women who had bought HC at least once between the ages of 15–55 (mean age at joining the EstBB (sd) = 35.6 (10.6), 32.8% below the age of 30). They contributed 1,040,298 person-years of follow-up (from the first purchase date), with an average follow-up of 14.2 years per person. Among all women using HC, 89.2% started using HC before joining the biobank, while 10.8% had their first purchase after joining. Their average age at the first HC purchase was 27.9 (sd = 9.2) years, with the highest number of initiators at the age of 19 (n_users_ = 4,842). The BMI of HC users during contraceptive use across all usage periods ranged from 14.1 to 43, with a mean of 24.3 (4.6). The distribution of BMI categories at the first prescription date showed that 61.8% (n_users_ = 45,136) of HC initiators were in the healthy weight category, 22.0% (n_users_ = 16,093) in the overweight, 11.9% (n_users_ = 8,683) in the obese, and 4.3% (n_users_ = 3,159) in the underweight category, which is comparable to BMI categories reported for the Estonian female population of similar age^53^.

Across nineteen years, twenty different HC formulations with five distinct routes of administration (intrauterine device, intravaginal ring, oral pill, transdermal patch and subdermal implant) were used. The most widely used HCs were COCs containing gestodene and ethinylestradiol (GSD/EE, n_users_ = 27,651) and drospirenone and ethinylestradiol (DRSP/EE, n_users_ = 26,030), levonorgestrel-releasing IUD (LNG IUD, n_users_ = 24,465), desogestrel-only pill (DSG, n_users_ = 16,557) and CHC ring containing etonogestrel and ethinylestradiol (n_users_ = 13,376). Detailed information about all purchased HCs across the study period is presented in Table 1.

**Table 1.**
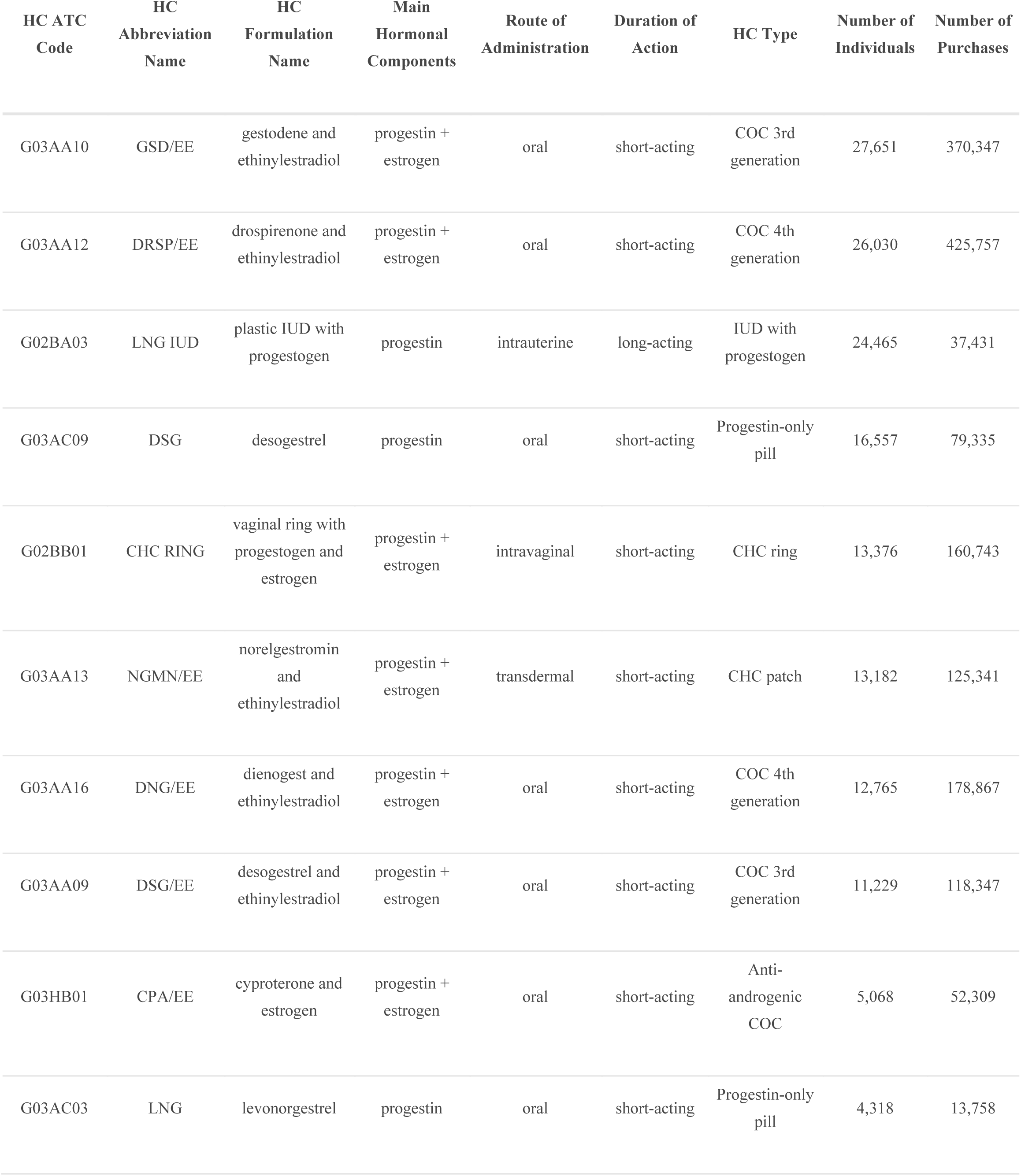
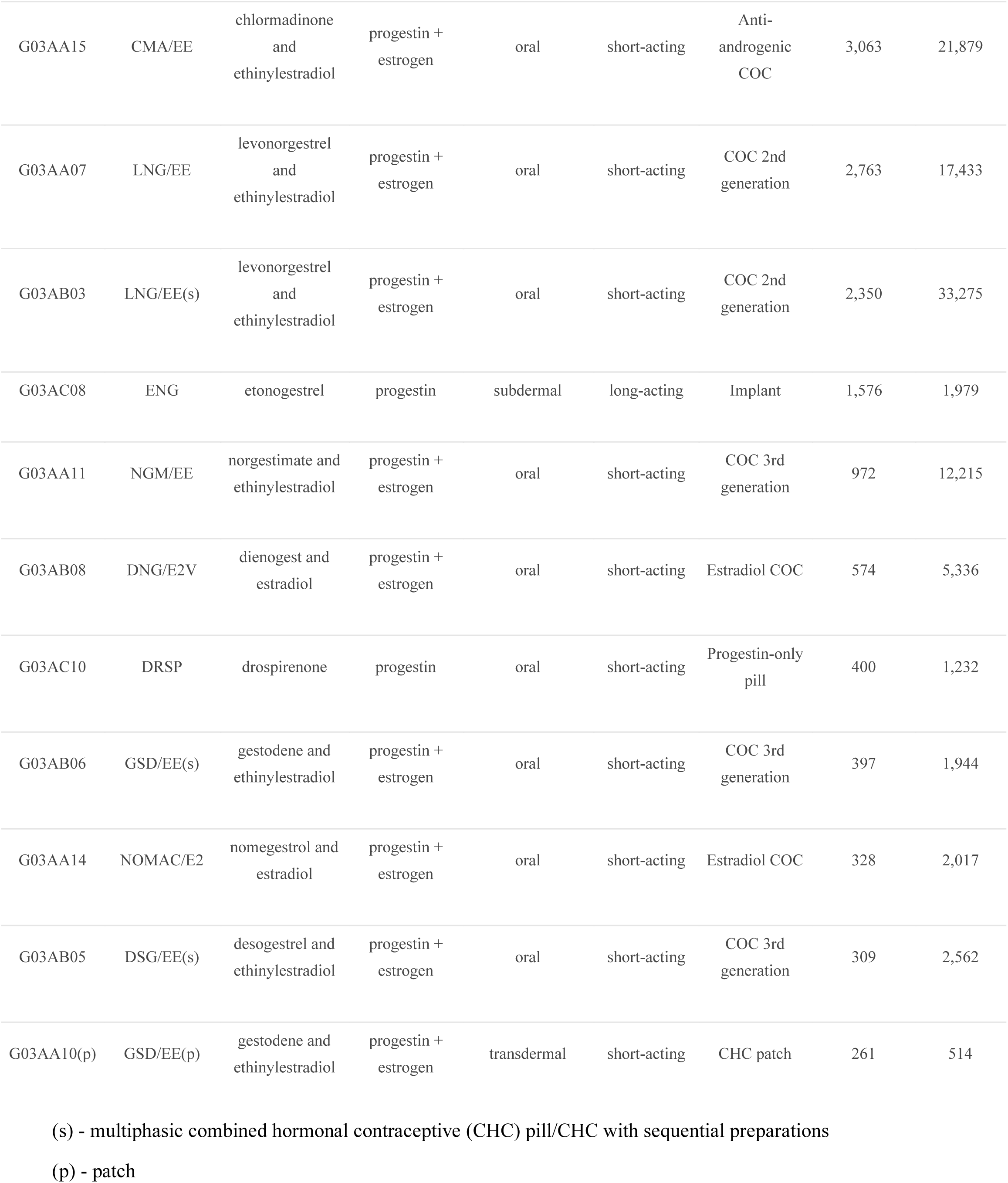
Overview of hormonal contraceptives (HCs) used by Estonian Biobank participants from 2004 to 2022, sorted by the total number of individuals using HCs.

The demographic structure of our study cohort has shifted significantly over the years, reflected in a notable decline in the proportion of individuals aged (constructed age, see Methods) 15–29, from 40.6% in 2004 to 15.3% in 2022 (Figure 1a, Supplementary Table 1). Accordingly, the annual prevalence rates of CHC users were much higher than POC users, but have decreased with time (Figure 1b, Supplementary Table 2), while prevalence rates of POC users steadily increased. Although our estimated prevalence rates were systematically higher than those reported in the study by Kurvits et al., who estimated annual prevalence rates of HC users aged 15–49 in Estonia from 2005 to 2019, the pattern of yearly changes was the same. Specifically, the annual prevalence rate of POC users in the EstBB increased across all age groups (15–19: from 0.3% in 2004 to 10.9% in 2022, 20–29: from 3.9% in 2004 to 17.2% in 2022, 30–39 from 3.2% in 2004 to 21.3% in 2022, 40–49: from 0.9% in 2004 to 27.6% in 2022 and 50–55: from 0.03% in 2004 to 18.0% in 2022). The annual prevalence rate of CHC users in the adolescent and the oldest age groups of EstBB increased (15–19: from 17.0% in 2004 to 38.2% in 2022, and 50–55: from 1.0% in 2004 to 2.0% in 2022), while for the other age groups it decreased (20–29 from 39.4% in 2004 to 26.6% in 2022, 30–39 from 21.5% in 2004 to 14.8% in 2022, and 40–49: from 9.6% in 2004 to 8.5% in 2022). Similar trends were observed by Kurvits et al., except our dataset showed an increase in CHC rates in groups 15–19 and 20–29 from 2015, and their study lacked information for the 50–55 group.

**Figure 1.**
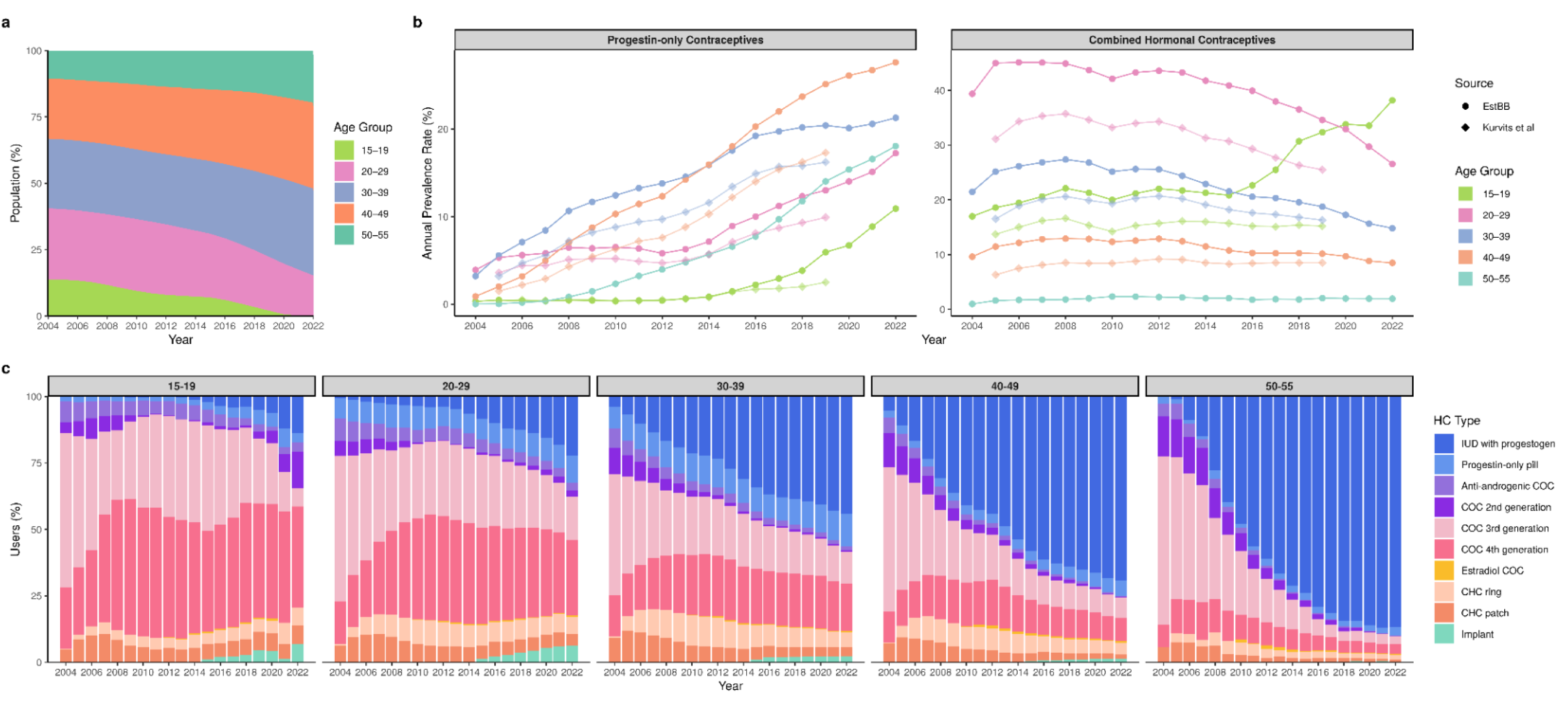
Overview of hormonal contraceptive (HC) use trends from 2004 through 2022. **a**: Age dynamics in the study sample (HC users and non-users combined) during the study period. **b**: Annual prevalence rate (%) of HC users in different age groups across studied years in Estonian Biobank (EstBB, darker colour) and Kurvits et al. (lighter colour), shown separately for two main HC classes - progestin-only contraceptives and combined hormonal contraceptives. **c**: Percentage of users of different HC types across studied years, stratified by age groups (15–19, 20–29, 30–39, 40–49, 50–55).

The changes in the percentage of different HC types used by women from different age groups throughout the study observation period are presented in Figure 1c and Supplementary Table 3. While generally there was a continuous decrease in anti-androgenic, 2nd and 3rd generation COC users, the proportion of 4th generation COC users was increasing, then remained steady in more recent years. An exception to this trend was found in the 15–19 and 20–29 age groups, where the proportion of 2nd generation COC users started increasing again from 2015. The use of estradiol COCs and HC implants began in 2010 and 2014, respectively. The proportions of different HC types varied slightly between years, reflecting the availability of specific formulations, as some multiphasic COCs and progestin-only pills (POPs), such as gestodene CHC (GSD/EE(s)), levonorgestrel-only pill (LNG) and drospirenone-only pill (DRSP), were only available on the market and used for 2–4 years.

While levonorgestrel-releasing IUDs (LNG-IUD) were increasingly used and primarily by older women (87% of users from the 50–55 age group in 2022), CHC dominated among younger and middle-aged women, especially in the early 2000s and 2010s, with the highest usage of 3rd and 4th generation COCs. On the other hand, POPs were most commonly used by the 20–29 and 30–39 age groups (10.2% and 12.2% in 2022, respectively), with these percentages remaining stable over the years. Estradiol COCs were the least used type of HCs and had minimal variation in the number of users across the studied period (up to 1.4% of users in all age groups throughout the years). CHC rings and patches were almost equally prevalent across all age groups and displayed a declining trend. Etonogestrel-releasing implants (ENG) were more common over the years among adolescents aged 15–19 (6.9% in 2022) and women aged 20–29 (6.4% in 2022), while among the users from older age groups, the proportions were steadily low.

### Hormonal contraceptive method initiation

The first-time prescriptions in 83.4% of women were CHCs, notably 36.1% being 3rd generation COCs and 24.9% 4th generation COCs, irrespective of their weight category (Supplementary Figure 3). However, each increase in the year of the first purchase was associated with lower percentages of 3rd generation COCs (decrease from 55.6% of initiators in 2004 to 14.8% in 2022) and higher percentages of LNG-IUDs (from 1.7% of initiators in 2004 to 36.0% in 2022). The 4th generation COCs peaked in 2011 with 38.7% of all initiators choosing this method. POPs were most used by the 30–39 age group (11.4%) and women from the obese weight category (9.4%), while LNG-IUDs were chosen by 39.5% of women from the 50–55 age group. Estradiol COC and ENG implant were the least used options for HC initiation (0.2% and 0.3%, respectively).

The first contraceptive period length of specific HC formulations varied from less than a month (<30 days) to more than three years (>1095 days), with a median of approximately 5.5 months (168 days) as the most common for the majority of HCs, except for long-acting methods, such as LNG-IUD and ENG implant, and multiphasic CHCs, such as LNG/EE(s) and GSD/EE(s) with higher medians (2190, 1027, 384, and 279 days, respectively, Figure 2). Some usage period lengths have been underestimated when the first purchase was made near the end of the observation period or when the woman continued using the method past the age of 55, when they exceeded the study age limits.

**Figure 2.**
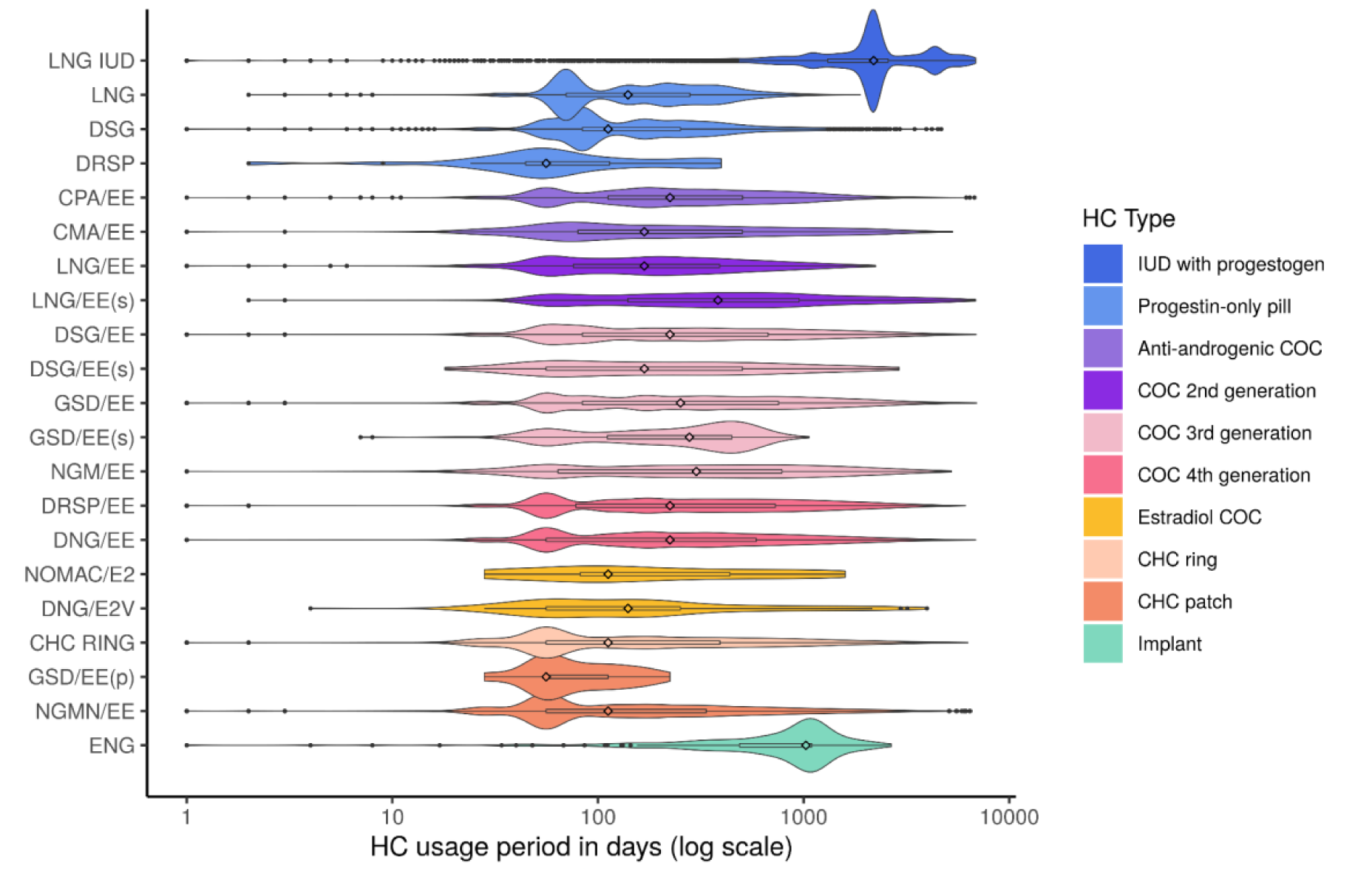
Length of first hormonal contraceptive (HC) usage period for different HC formulations. Specific HC formulations are listed on the y-axis, while the HC formulation usage period length in days (days shown on the logarithmic scale) is plotted along the x-axis. The colour illustrates the HC type. The central diamond shapes mark the mid-point value, and the black dots mark the outliers.

Looking into the ICD-10 three-character category diagnosis codes recorded up to three months before the first HC usage period start date, we found 1,233 different codes, where 53.1% (n_users_ = 38,820) of HC initiators had Z30 diagnosis code related to contraceptive management and 11.2% (n_users_ = 8151) did not have any record of the medical condition. Additionally, 4.8% (n_users_ = 3531) had heavy menstrual bleeding (N92), 4.0% (n_users_ = 2922) medical abortion (O04), 2.4% (n_users_ = 1744) dysmenorrhoea (N94), 1.6% (n_users_ = 1202) acne (L70), and 0.7% (n_users_ = 472) PCOS (E28.2), which were from the list of curated diagnoses we considered as a potential reason for starting HCs.

Previous studies have shown changes in prescribing and purchasing trends during the COVID-19 pandemic due to lockdowns and limited access to healthcare services^54^. We investigated the impact of the COVID-19 pandemic and pandemic-induced lockdowns on HC initiation, comparing 2019 (pre-COVID-19 period) to 2020 and 2021 (COVID-19 periods). Overall, up to 55.4% of first-ever initiators were consistently from the 20–29 age group, and up to 50.0% of initiators with previous HC use history were from the 30–39 age group. Our longitudinal cohort’s yearly number of HC users gradually declined from 2019 to 2021. While the population-weighted average rate of first-ever initiators decreased from 1.3 to 1.1 to 0.8 during this period, other initiators decreased from 9.4 to 8.6 to 8.3, which likely also reflects the ageing dynamics of our study cohort. Furthermore, the overall monthly purchase trends differed notably for 2020 (Supplementary Figure 4a), where March 2020 was 20% above the grand mean and April 2020 was 20% below it. This sharp drop in HC purchases from March 2020 to April 2020, with recovery beginning in June 2020, aligned with the first restrictions in Estonia that started on 16th March 2020, which gradually eased by June 2020. Specifically, March had higher purchase numbers of CHC pills, patches, rings, and POPs than March 2019, likely due to the stockpiling behaviour, with the peak on March 13th (Supplementary Figure 4d), when the Estonian government announced a state of emergency. Conversely, LNG-IUDs and implants had their lowest record numbers in March and April 2020. Following June 2020, only 2nd generation COCs and LNG-IUDs had a slight increase in purchases compared to the corresponding periods in 2019. Also, compared to 2021, only 2nd generation and estradiol COCs, implants, LNG-IUDs and POPs returned to similar numbers as recorded in the pre-COVID-19 period.

### Contraceptive method switching

While 35.7% (n_users_ = 26,109) of women used only one drug formulation during the observed study period, 64.3% (n_users_ = 46,962) switched to a different one (ranging between 2 to 11 different HC formulations, Figure 3a). Specifically, out of all HC users, 23.9% (n_users_ = 17,453) were classified as non-switchers, 46.5% (n_users_ = 33,991) as broad switchers, 17.7% (n_users_ = 12,929) as rapid switchers, 8.7% (n_users_ = 6,346) as rapid discontinuers (7.3% (n_users_ = 5,362) with only one usage period and 1.4% (n_users_ = 984) with several usage periods), while 3.2% (n_users_ = 2,352) did not fit into any of the defined categories. The latter group included, for example, users of one HC formulation for less than 180 days, but more than 90 days (see Methods). From these types of users, we were interested in non-switchers, rapid switchers and rapid discontinuers with only one usage period (user characteristics presented in Supplementary Table 4). Non-switchers and rapid switchers were predominantly women from the 20– 29 age group (33.84% and 49.1%, respectively), while rapid discontinuers were from the 30–39 age group (32.2%). The non-switcher usage periods ranged from 182 to 6,934 days, with an average of 1,658 days (corresponding to 4.5 years), while in rapid switchers, the average time to switch from one formulation to a different one was 49 days.

**Figure 3.**
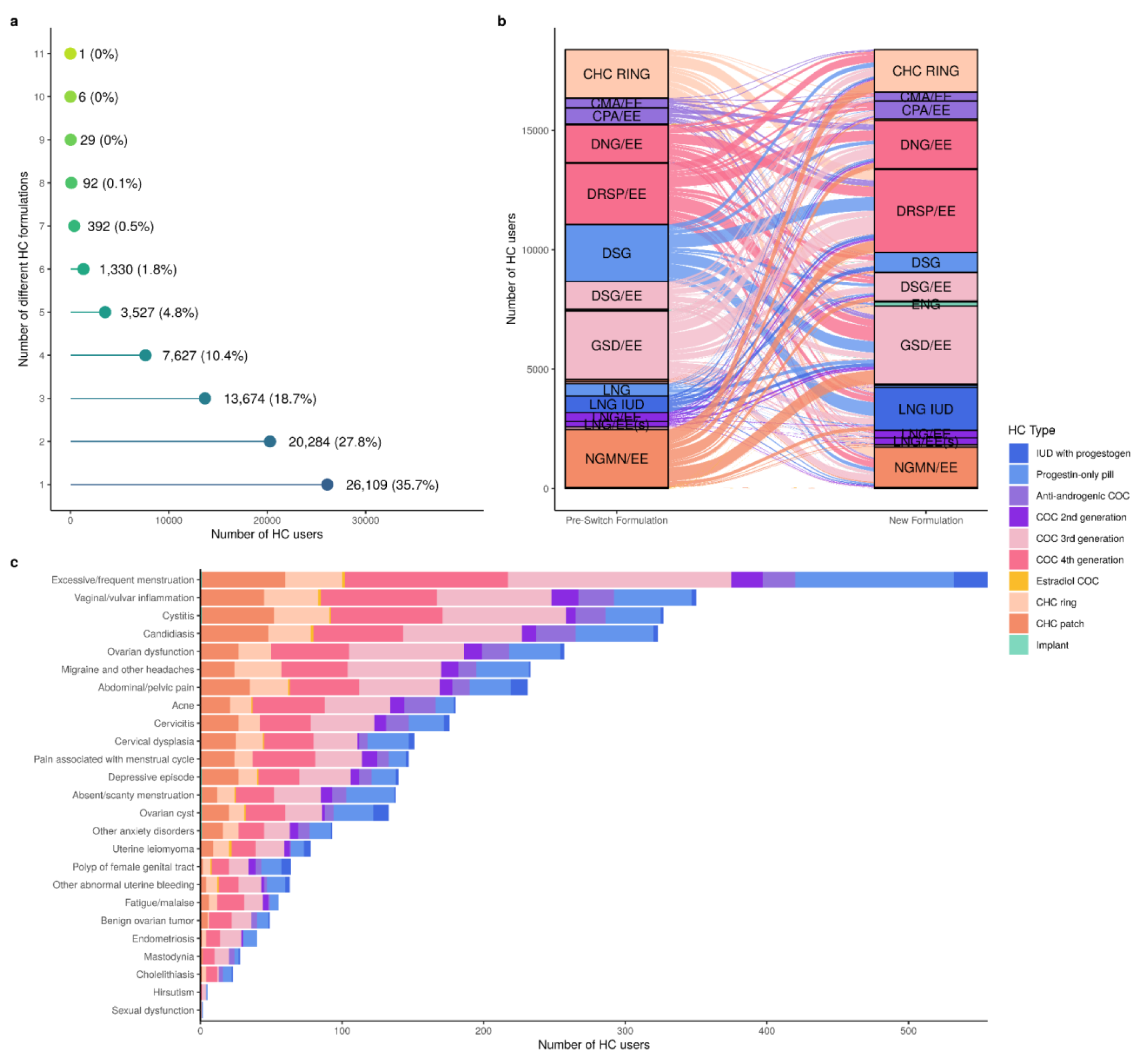
Hormonal contraceptive (HC) usage behaviour. **a**: Number of different HCs purchased per individual during the study observation period. **b**: Graphical illustration of switching pathways of rapid switchers (pre-switch formulation is the HC method individuals switched from, while the new formulation is the HC method individuals switched to). The colour illustrates the HC type. **c**: Main ICD-10 diagnoses recorded between the first purchase of pre-switch HC formulation and the new formulation in rapid switchers. The coloured bars indicate the type of pre-switch HC method.

The illustration of the HC switching behaviour of rapid switchers with complex multidirectional flows is presented in Figure 3b. The HC formulations with the highest number of switches were short-acting HCs with the highest usage volume, such as GSD/EE, DRSP/EE, norelgestromin and ethinylestradiol (NGMN/EE) patch, DSG and CHC ring, each with 18–22% of rapid switchers and around 10% of drug users, except 18.4% of NGMN/EE users, and 14–15% of DSG and CHC ring users. The subsequent HC formulations for these switchers were similar and overlapping, where GSD/EE rapid switchers mostly switched to DRSP/EE (25.2%), DNG/EE (14.9%), and NGMN/EE patch (10.9%), DRSP/EE users switched to GSD/EE (23.5%), DNG/EE (16.6%), and CHC ring (13.1%), and NGMN/EE users switched to GSD/EE (23.8%), CHC ring (21.0%) and DRSP/EE (19.6%). Most notably, switching to DNG/EE, LNG-IUD, and ENG implant were more common than switching from them. Among rapid switchers who switched from COCs (n_users_ = 7,800 (60.3%)), 66.5% switched to a different COC, 11.3% switched to a CHC patch, and 11.1% to the CHC ring. Those who switched to the ring used the new method for approximately 4.9 months (median value) compared to the 3.7 months for a patch.

In the period between the first purchase date of the pre-switch HC formulation and the first purchase date of the new formulation, no ICD-10 diagnosis codes were identified for 50.2% (n_users_ = 6,486) of rapid switchers, while 23.1% (n_users_ = 2,982) had diagnoses from our curated list of potential side effects (see Methods). Notably, we observed diagnoses related to reproductive system disorders (e.g. excessive/frequent menstruation (n_users_ = 556)), neurological/pain conditions (migraine and other headaches (n_users_ = 233), acne (n_users_ = 180), menstrual pain (n_users_ = 147)), mood disorders (depressive episode (n_users_ = 140), anxiety (n_users_ = 93)), and mastodynia (n_users_ = 28) (Figure 3c, Supplementary Table 5). In addition to this, HC switching could be due to the contraceptive failure leading to abortion^55^. In our cohort, medical abortion (ICD-10 O04) overlapped with inferred HC usage periods in 3.2% (n_users_ = 2,359, n_periods_ = 2,440) of all users, with COCs being the most commonly used contraceptive before abortion. The majority of these women (77.3% (n_users_ = 1,823)) used HCs again (median time to the new HC formulation = 684 days/1.9 years (11–6,132 days)), where other COCs (n_periods_ = 792 (32.5%)), LNG-IUD (n_periods_ = 125 (5.1%)) and POPs (n_periods_ = 122 (5.0%)) were the most common choices following abortion. Similarly, in 2.2% (n_rapid_ = 288, n_periods_ = 291) of rapid switchers, abortion diagnosis was recorded between purchases of two HC formulations. The most common switches were from one short-acting method to another of comparable effectiveness and safety, such as from one COC to a different COC (n_periods_ = 79 (27.1%)), from a CHC patch to a COC (n_periods_ = 43 (14.7%)), and from a COC to the CHC ring (n_periods_ = 28 (9.6%)).

### Presence of thromboembolism risk factors

Among 13,610 HC users with purchases in 2022, which is the most recent year with complete data, 63.7% (n_users_ = 9,052) were CHC users (mean age at purchase (sd) = 33.9 (8.0)) and 36.3% (n_users_ = 5,148) POC users (mean age at purchase = 36.8 (7.9)). Thromboembolism was diagnosed in 43 women (VTE/ATE diagnosis recorded after the first HC purchase date in 2022; mean age at diagnosis = 39.8 (7.4)), and out of those, 38 were diagnosed during the inferred HC usage period. The investigation of thromboembolism risk factors (Figure 4) revealed their higher prevalence among POC users (at least one risk factor) than among CHC users. The most common risk factors were age above 35 (54.4% of POC users and 40.6% of CHC users), purchase of risk-increasing drugs (14.8% of POC and 17% of CHC), obesity (14.6% of POC and 10.1% of CHC), and migraine (8.6% of POC and 5.6% of CHC). The percentage for carrier status of thrombosis risk-increasing genetic variants was slightly higher among POC users (5.6% of POC vs 4.7% of CHC), while the difference for personal history of VTE/ATE (3.4% of POC and 1.8% of CHC) was almost 2-fold. The differences in family history of thrombosis, other thrombosis risk conditions and pneumonia between POC and CHC users were not statistically significant (P > 0.05). Out of 13,610 analysed HC users, 61.7% (n_users_ = 8,397) had at least one first-degree relative in the EstBB, and 7.4% (n_users_ = 1,022) had a first-degree relative diagnosed with venous or arterial thromboembolism before HC purchase. Women using HCs who had at least one first-degree relative diagnosed with thromboembolism had 1.7-fold higher odds of being a FVL or PTM carrier (76/946 (8.0%) versus 327/7,048 (4.6%), Fisher’s exact test P < 0.001). The sensitivity and specificity of family history of thrombosis for identifying carrier status were 18.9% (95% CI 15.0–22.7%) and 88.2% (95% CI 87.5–88.9%), respectively.

**Figure 4.**
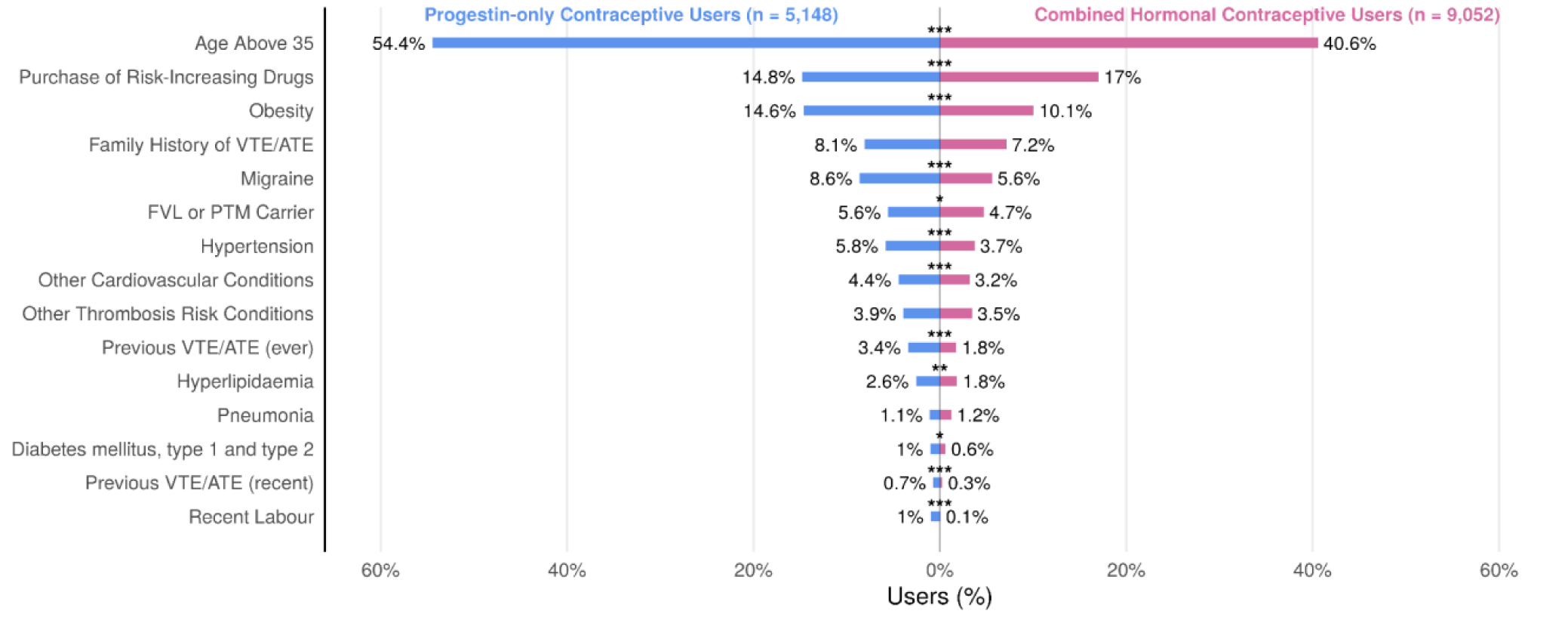
Thromboembolism risk factors among hormonal contraceptive (HC) users with purchases in 2022, displayed separately for the progestin-only contraceptive group and the combined hormonal contraceptive group. On the y-axis are risk factors, while on the x-axis are the numbers of HC users and within-group percentages. The asterisk above the bar marks the level of significance from Fisher’s exact test (P < 0.05 ∼ “*”, P < 0.01 ∼ “**”, P < 0.001 ∼ “***”).

Across the entire study group, we found that 5.3% (n_users_ = 3,886) of HC users carried at least one VTE high-risk genetic variant (3.6% (n_users_ = 2,610) only FVL, 1.8% (n_users_ = 1,330) only PTM, 0.07% (n_users_ = 54) both FVL and PTM). While in 1.5% (n_users_ = 1,060) of users, thrombosis diagnosis occurred during the inferred HC usage period, of those, only 17.6% purchased antithrombotic agents (ATC group B01A) after the diagnosis and while HCs were in active use according to purchasing data. The top five ICD-10 thrombosis codes were I80 (unspecified phlebitis and thrombophlebitis, n_users_ = 330), I80.2 (deep vein thrombosis, n_users_ = 212), I26.9 (pulmonary embolism, n_users_ = 130), I80.3 (embolism or thrombosis of lower extremity, n_users_ = 125) and I82.8 (embolism and thrombosis of vena cava, n_users_ = 78). Moreover, 8.6% of these thrombosis cases carried either the FVL or PTM variant. Overall, we observed a significantly higher percentage of thrombosis cases at any time point in carriers using HC (6.5%) compared to the non-carriers (4.2%; χ² = 49.61, df = 1, P < 0.001).

## Discussion

In this study, leveraging EstBB data and HC purchasing data from the period 2004 to 2022, we aimed to describe in detail HC use trends of >73,000 EstBB female participants, compare results with the Estonian population HC use trends and evaluate the suitability of longitudinal biobanks, such as EstBB, for studying genetic risk for HC side effects. Our described use trajectories of various HCs were generally consistent with the population-level statistics, albeit with slight differences discussed below. We identified several HC user types, such as non-switchers, broad switchers, rapid switchers, and rapid discontinuers, with only one or several usage periods. While 64.3% of women using HC eventually switched to a different formulation, 17.7% switched within 3 months of the first purchase, indicating potential side effects^7–9^. Moreover, we detected first-time records of ICD-10 diagnosis codes related to potential HC side effects in 23.1% of rapid switchers, with heavy menstrual bleeding being the most common. We also estimated the prevalence of genetic variants associated with higher VTE risk in 5.3% of women using HC and evaluated various arterial and venous thromboembolism risk factors, including personal and family history of thrombosis.

Compared with Estonian national statistics^18^, we had slightly higher POCs and CHCs prevalence rates. This could be due to the set maximum length for long-acting methods, including CPA/EE HC, and incorporating short pauses (<90 days between two purchases) as part of the usage period. Also, EstBB 15–29 age groups have drastically reduced in size in recent years; therefore, the prevalence rates became “artificially” bigger, as no new participants were recruited into EstBB after 2019. Regarding higher implant use among younger women, increased POC use in recent years, the presence of the most common risk factors and POC/CHC use among women with risk factors, our results are in line with those from Kurvits et al. The slight differences in risk factor percentages between the studies can be explained by our additional use of BMI values to define obesity and the inclusion of the 50–55 age group. In addition, leveraging genetic data and genetically inferred relatives among EstBB participants, we found that the family history of thrombosis did not significantly differ between POC and CHC users, although it is important to note that not all women had first-degree relatives in the EstBB. Finally, we observed that some women continued to purchase HCs even after the thrombosis diagnosis, without the concomitant purchase of antithrombotic agents^56^. However, this finding should be interpreted cautiously, since drug purchase data does not reflect medications administered during inpatient care.

Several observational studies evaluating contraceptive use in Nordic countries and the UK reported increasing use of levonorgestrel-containing COCs, POPs and IUDs^57–60^. However, in EstBB, we only see an increased use of IUDs, which aligns with observations from the Estonian overall population^18^. Next, a study investigating prescription rates of contraceptives during the COVID-19 pandemic in England discovered an overall significant decrease in prescriptions, and an increase in POPs prescriptions^54^, which might be relevant to consider when analysing use trends during this period. While we focused on purchasing and not prescription dates, we observed a higher number of CHC and POP purchases right before lockdown and a lower number during the first year of the COVID-19 pandemic, especially of implants and LNG-IUDs. However, these differences were not statistically significant. Estonia has a well-organised and digitalised healthcare system, and there has never been a complete lockdown. Also, the ageing cohort could partially explain the gradual decrease in HC initiators; while the number of women using HCs globally is growing as the population rises and renews^1^, the number of women at peak reproductive age in the EstBB is decreasing.

While population-based studies are great for describing use trends and general risk factors, they generally lack genetic data for evaluating the presence of specific large-effect genetic risk factors. Here, population-based biobanks with readily available genetic data have an advantage. It has been shown that up to 20% of individuals diagnosed with VTE (25% in the case of idiopathic VTE) carry at least one risk-increasing genetic variant^26^. We found that 5.3% of women using HC carried either FVL or PTM variant, and had a significantly higher percentage of thrombosis diagnoses than non-carriers. At the same time, generally, only family history is evaluated before HC prescription, serving traditionally as a proxy for genetic risk. However, family history of thrombosis does not always adequately reflect carrier status^61,62^. Readily available information on carrier status in biobanks can potentially reduce the incidence of VTE among women on HC. In parallel, the stratification based on venous and arterial thrombosis polygenic risk scores could possibly offer a more comprehensive approach to identifying women requiring more careful contraceptive selection^29^ and eligible for additional thrombophilia screening or nAPCsr blood coagulation testing.

When it comes to large-scale genetic analyses of drug use and associated side effects, defining cases and controls is the first step, but, unlike for cardiometabolic drugs with clear physiological endpoints, doing so for hormonal medications can be challenging. Using a similar approach from Lo et al.^63^, but contextually adjusted, we identified several HC user types, such as non-switchers, broad switchers, rapid switchers, rapid discontinuers with only one usage period and rapid discontinuers with several usage periods that could be used as phenotypes of interest. In general, switching between different HC formulations is a common part of women’s contraceptive journey throughout their reproductive years^7^. However, rapid discontinuation or switching to a different drug formulation can be due to the side effects^7–9^. For that reason, we focused further on rapid switchers and investigated ICD-10 diagnoses recorded between the first purchase of pre-switch HC formulation and the first purchase of post-switch HC formulation. Indeed, we found records of diagnoses related to potential side effects. The observed prevalence of these diagnoses among only 23.1% of rapid switchers could potentially be explained by underreporting or other causes of switching, and a different study design would be needed to clarify these. Similarly, from inferred usage periods of first-ever initiators, we observed the highest number of women on short-acting methods clustered around 90 days period length. This coincides with the general recommendation to use HCs for at least three months to allow the body to adapt to exogenous hormones^64,65^, but it also suggests that for many women, initial HC method matching is suboptimal. This highlights the need for developing more effective approaches to HC selection, potentially accounting for individual genetic risk as a modifier of side effects associated with HC use.

The key strength of our study is quality EstBB datasets derived from multiple sources, including genetic and detailed clinical information, that ensure comprehensive analysis of individual HC use over a time period longer than 10 years. Additionally, our data and conducted analyses are free from self-report and recall bias, which was shown to increase errors and reduce research reproducibility significantly^66,67^. As far as we are aware, this is the first study to assess HC use based on purchase data within a biobank setting.

Our study has several limitations that need to be mentioned. First, the described HC trends are specific to Estonia, and the investigated individuals are of European ancestry, which may not apply to other healthcare systems and ancestries. Second, the inferred HC usage periods from the purchase data may not accurately represent actual usage and adherence to the HC regime. This may lead to a slight overestimation of the usage periods. However, the short-acting HC effects extend beyond inconsistent adherence due to the gradual adaptation of the body to the hypothalamic-pituitary-gonadal axis natural rhythm after discontinuation. Third, we could not reliably track the removal of the long-term acting contraceptives with the ICD-10 coding system; we were able to adjust only using pregnancy dates. The distinction of separate four-character subcategory codes for IUD and implant insertion, checking, and removal by the medical community would benefit future observational studies. Fourth, we did not have access to the information on purchases of emergency contraceptives and injections. Emergency contraceptives are “over-the-counter” drugs in Estonia, and our input was prescription purchases and refills. The injectable medroxyprogesterone is officially an unauthorised product, marketed only with a special permit from the Estonian Agency of Medicines and used only on the specific doctor’s recommendation; therefore, we anticipated a negligible number of users. Fifth, given the current research setting, which is different from the gold standard Phase 3 clinical studies, we could not know certain reasons for switching between different formulations, nor whether the discontinuation is due to some side effect or wish to become pregnant. Future implementations of free-text data mining of unstructured EHRs and epicrises could partially aid in mitigating this limitation. Sixth, we acknowledge that the presented numbers are just study population averages, while HC research requires more personalised insights^68^. However, our valuable cohort-level statistic, compared to the national-level statistic, provides a baseline that will enable meaningful interpretation of individual variations. At the same time, population-based biobanks facilitate future personalised approaches to HC selection.

In conclusion, our insights into real-world HC purchases of EstBB participants, inferred usage periods, and HC switching offer additional context and support for understanding women’s preferences in HCs, as well as the safety of HCs and effectiveness during typical use. Finally, this study can serve as a foundation for exploring different research questions of HC use, including genetic and microbiome data from the population-based biobanks for robust large-scale genetic analyses as a step towards more personalised HC use, reducing the number of individuals experiencing harmful and unpleasant side effects.

## Data Availability

Pseudonymized data and/or biological samples can be accessed for research and development purposes in accordance with the Estonian Human Genome Research Act (https://www.riigiteataja.ee/en/eli/ee/531102013003/consolide/current). To access data, the research proposal must be approved by the Scientific Advisory Committee of the Estonian Biobank as well as by the Estonian Committee on Bioethics and Human Research. Access to samples requires the same approval process and an additional approval from the Senate of the University of Tartu. For more details on data access and relevant documents, please see https://genomics.ut.ee/en/content/estonian-biobank#dataaccess.

## Acknowledgements

The authors thank the Estonian Biobank participants for their contribution to this work, and the Estonian Biobank research team (Andres Metspalu, Lili Milani, Tõnu Esko, Reedik Mägi, Mait Metspalu, Mari Nelis and Georgi Hudjashov) for data collection, genotyping, QC and imputation. We also thank the High-Performance Computing Center of the University of Tartu for providing computational resources for data analysis.

## Author Contributions

T.L. conceived the study. J.D. performed the analysis, interpreted the results and wrote the first draft.

T.L. and M.M. contributed to the study design, data analysis and data interpretation. T.L., M.M., H.C., K.L., M.E., R.M., and L.M. provided critical review and supervision. All authors read and approved the final manuscript.

