## Supplementary for "Hormonal contraceptive drug use trends in the Estonian Biobank"

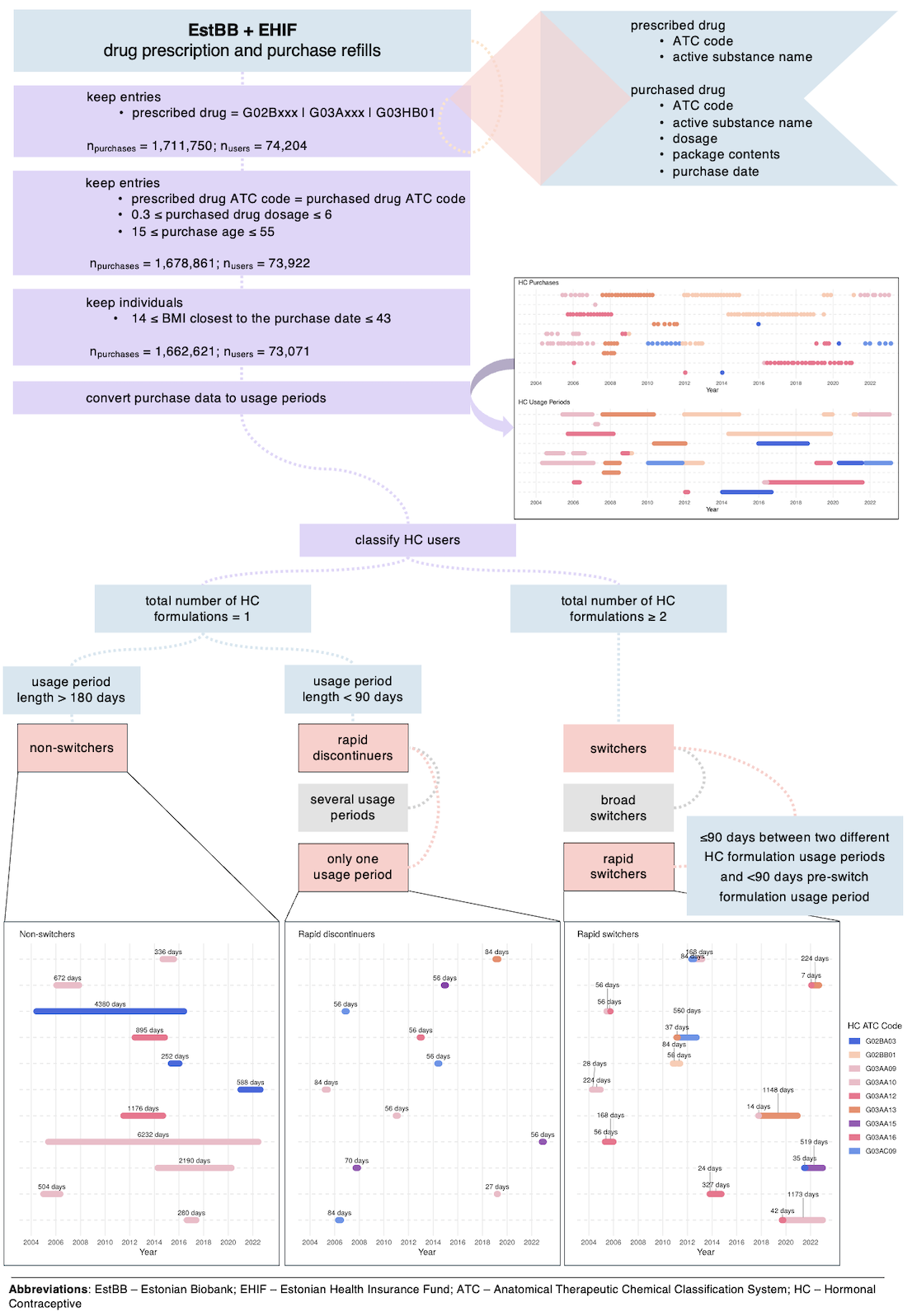

**Supplementary Figure 1.** Illustration of the pipeline for defining hormonal contraceptive (HC) usage periods from HC purchase data and classification of different HC user types. The data points from the figure are for illustrative purposes and are not real observations.

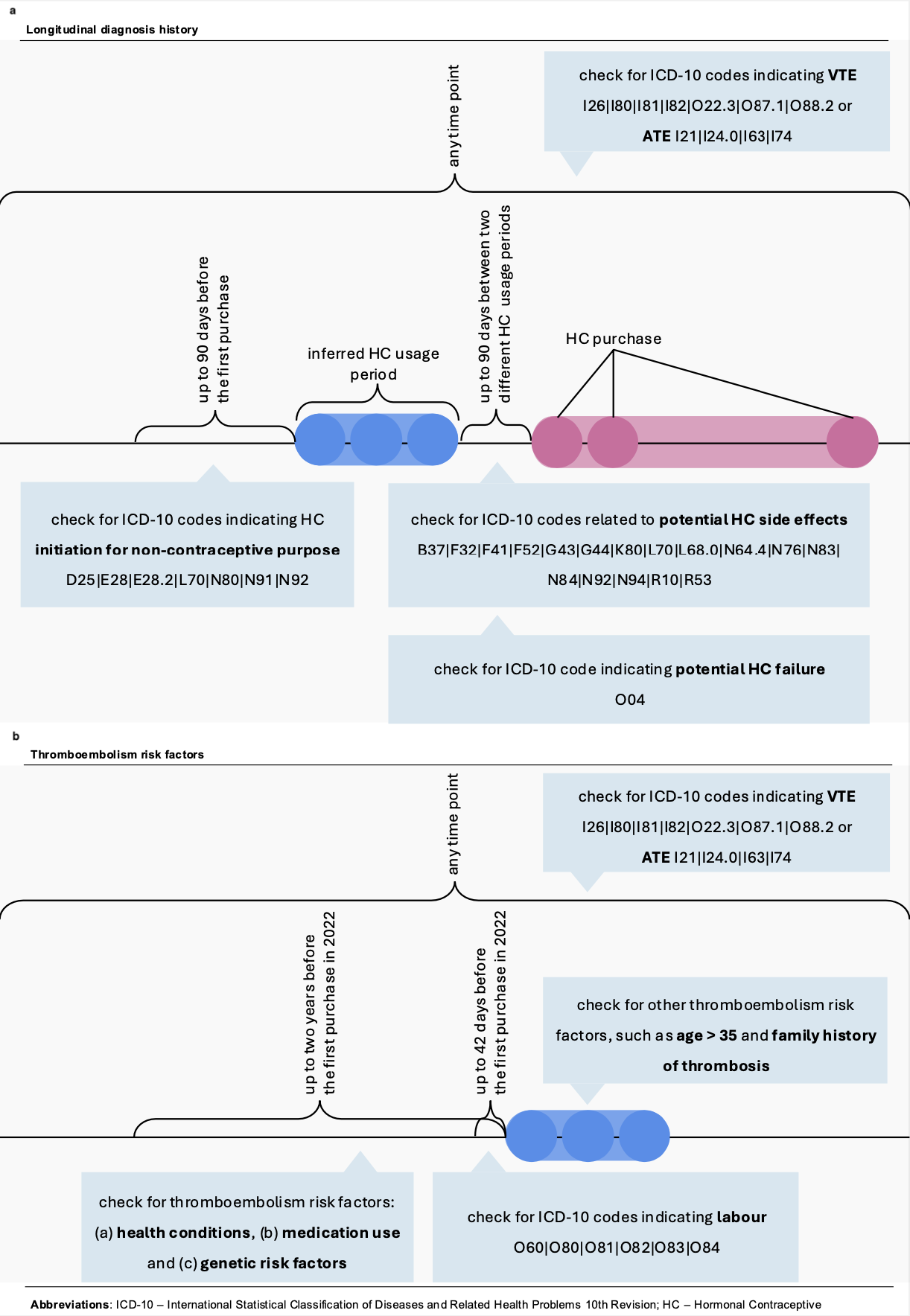

**Supplementary Figure 2.** Illustration of the strategy used to identify relevant ICD-10 diagnosis codes and thromboembolism risk factors before, during and after hormonal contraceptive (HC) use. **a**: identification of ICD-10 diagnosis codes. **b**: identification of thromboembolism risk factors.

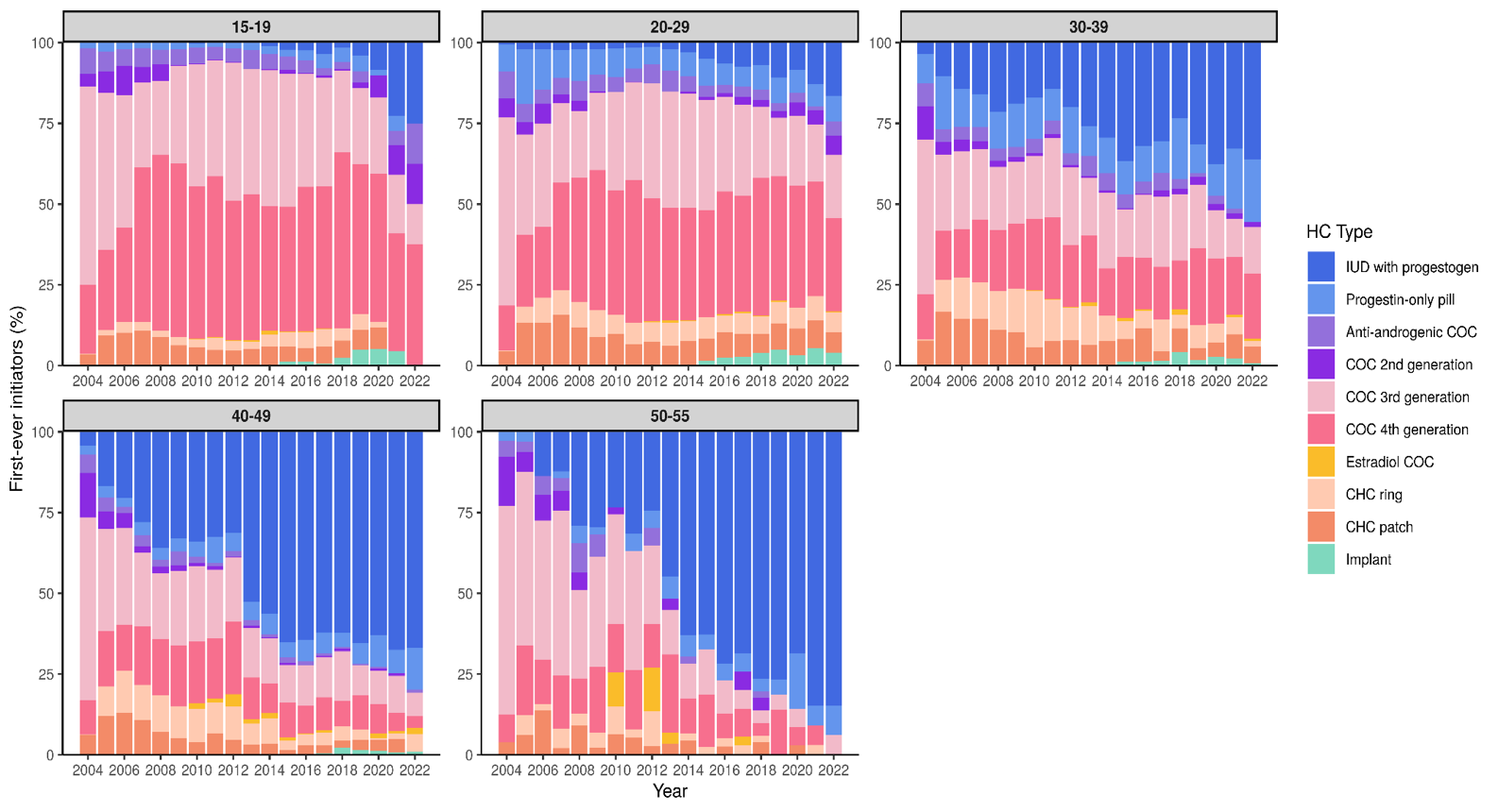

**Supplementary Figure 3.** The percentage of different hormonal contraceptive (HC) types of initiators (first-ever observed HC usage periods in study cohort) across studied years, stratified by age groups (15–19, 20–29, 30–39, 40–49, 50–55).

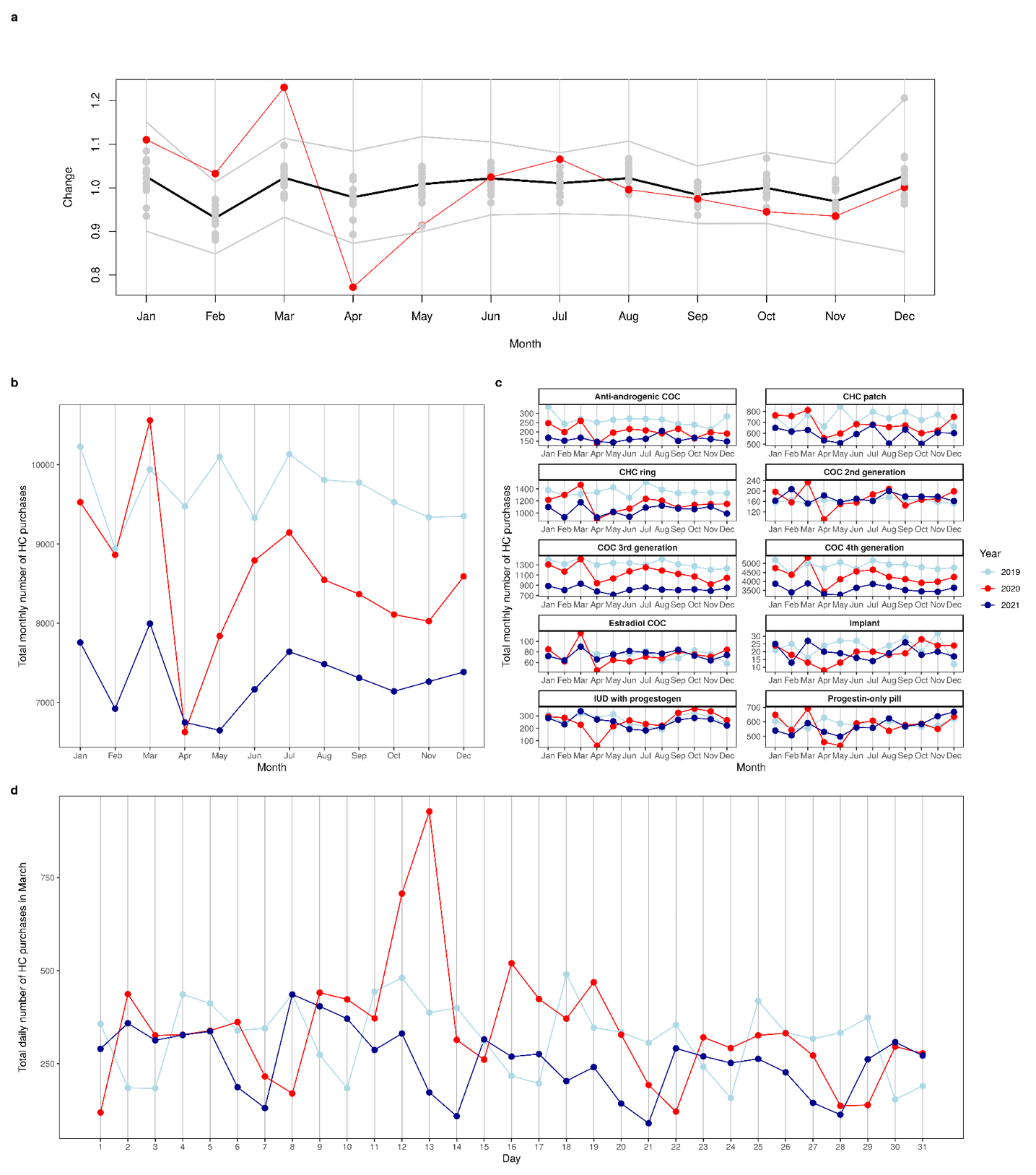
**Supplementary Figure 4.** Hormonal contraceptive (HC) purchase trends in 2019 (pre-COVID-19 period), 2020 and 2021 (COVID-19 periods). **a**: Monthly deviation ratio from the yearly average for years 2005 through 2021 (exact value marked with the dot). The red colour and line mark 2020, while the black and grey lines mark the mean across all years and ±3 standard deviations, respectively. **b**: Comparison of the total number of HC purchases per month in the years 2019 (light blue line), 2020 (red line) and 2021 (dark blue line). **c**: The total number of monthly HC purchases in 2019, 2020 and 2021, stratified by HC type. **d**: The total daily number of HC purchases in March 2019, 2020 and 2021.

**Supplementary Table 1.** Total number of individuals from specific age group in the whole study cohort (hormonal contraceptive (HC) users and HC non-users)

|  | **age_group** | | | | | |
| --- | --- | --- | --- | --- | --- | --- |
| **year** | **15-19** | **20-29** | **30-39** | **40-49** | **50-55** | **15-55** |
| 2004 | 13607 | 26268 | 25730 | 22254 | 10398 | 98257 |
| 2005 | 13644 | 26352 | 25999 | 22599 | 10646 | 99240 |
| 2006 | 13360 | 26418 | 26255 | 22832 | 11121 | 99986 |
| 2007 | 12776 | 26485 | 26466 | 23154 | 11542 | 100423 |
| 2008 | 11750 | 26753 | 26452 | 23571 | 11895 | 100421 |
| 2009 | 10568 | 26992 | 26402 | 24014 | 12345 | 100321 |
| 2010 | 9460 | 27057 | 26233 | 24526 | 12689 | 99965 |
| 2011 | 8579 | 26785 | 26109 | 24811 | 13173 | 99457 |
| 2012 | 7884 | 26251 | 26055 | 25133 | 13484 | 98807 |
| 2013 | 7481 | 25216 | 26205 | 25467 | 13657 | 98026 |
| 2014 | 7136 | 24172 | 26254 | 25690 | 13970 | 97222 |
| 2015 | 6744 | 23100 | 26332 | 25958 | 14072 | 96206 |
| 2016 | 5704 | 21935 | 26401 | 26209 | 14101 | 94350 |
| 2017 | 4369 | 20657 | 26463 | 26425 | 14094 | 92008 |
| 2018 | 3075 | 19230 | 26729 | 26411 | 14224 | 89669 |
| 2019 | 1768 | 17704 | 26966 | 26362 | 14573 | 87373 |
| 2020 | 580 | 16201 | 27029 | 26183 | 14924 | 84917 |
| 2021 | 158 | 14279 | 26754 | 26053 | 15346 | 82590 |
| 2022 | 55 | 12245 | 26222 | 25979 | 15744 | 80245 |

**Supplementary Table 2.** Annual prevalence rates of hormonal contraceptive (HC) users in the Estonian Biobank (EstBB) and the Estonian population (Kurvits et al. (2021)^1^)

| **hormonal_composition** | **age_group** | **year** | **prevalence_rate** | **source** |
| --- | --- | --- | --- | --- |
| progestin | 15-19 | 2004 | 0.32 | estbb |
| progestin + estrogen | 15-19 | 2004 | 16.97 | estbb |
| progestin | 20-29 | 2004 | 3.90 | estbb |
| progestin + estrogen | 20-29 | 2004 | 39.36 | estbb |
| progestin | 30-39 | 2004 | 3.21 | estbb |
| progestin + estrogen | 30-39 | 2004 | 21.46 | estbb |
| progestin | 40-49 | 2004 | 0.89 | estbb |
| progestin + estrogen | 40-49 | 2004 | 9.61 | estbb |
| progestin | 50-55 | 2004 | 0.03 | estbb |
| progestin + estrogen | 50-55 | 2004 | 0.98 | estbb |
| progestin | 15-19 | 2005 | 0.48 | estbb |
| progestin + estrogen | 15-19 | 2005 | 18.58 | estbb |
| progestin | 15-19 | 2005 | 0.40 | kurvits |
| progestin + estrogen | 15-19 | 2005 | 13.70 | kurvits |
| progestin | 20-29 | 2005 | 5.32 | estbb |
| progestin + estrogen | 20-29 | 2005 | 44.98 | estbb |
| progestin | 20-29 | 2005 | 3.60 | kurvits |
| progestin + estrogen | 20-29 | 2005 | 31.10 | kurvits |
| progestin | 30-39 | 2005 | 5.56 | estbb |
| progestin + estrogen | 30-39 | 2005 | 25.15 | estbb |
| progestin | 30-39 | 2005 | 3.20 | kurvits |
| progestin + estrogen | 30-39 | 2005 | 16.50 | kurvits |
| progestin | 40-49 | 2005 | 2.01 | estbb |
| progestin + estrogen | 40-49 | 2005 | 11.45 | estbb |
| progestin | 40-49 | 2005 | 1.50 | kurvits |
| progestin + estrogen | 40-49 | 2005 | 6.30 | kurvits |
| progestin | 50-55 | 2005 | 0.05 | estbb |
| progestin + estrogen | 50-55 | 2005 | 1.61 | estbb |
| progestin | 15-19 | 2006 | 0.39 | estbb |
| progestin + estrogen | 15-19 | 2006 | 19.44 | estbb |
| progestin | 15-19 | 2006 | 0.50 | kurvits |
| progestin + estrogen | 15-19 | 2006 | 15.00 | kurvits |
| progestin | 20-29 | 2006 | 5.60 | estbb |
| progestin + estrogen | 20-29 | 2006 | 45.11 | estbb |
| progestin | 20-29 | 2006 | 4.40 | kurvits |
| progestin + estrogen | 20-29 | 2006 | 34.30 | kurvits |
| progestin | 30-39 | 2006 | 7.09 | estbb |
| progestin + estrogen | 30-39 | 2006 | 26.14 | estbb |
| progestin | 30-39 | 2006 | 4.70 | kurvits |
| progestin + estrogen | 30-39 | 2006 | 18.90 | kurvits |
| progestin | 40-49 | 2006 | 3.18 | estbb |
| progestin + estrogen | 40-49 | 2006 | 12.15 | estbb |
| progestin | 40-49 | 2006 | 2.20 | kurvits |
| progestin + estrogen | 40-49 | 2006 | 7.50 | kurvits |
| progestin | 50-55 | 2006 | 0.20 | estbb |
| progestin + estrogen | 50-55 | 2006 | 1.71 | estbb |
| progestin | 15-19 | 2007 | 0.38 | estbb |
| progestin + estrogen | 15-19 | 2007 | 20.61 | estbb |
| progestin | 15-19 | 2007 | 0.40 | kurvits |
| progestin + estrogen | 15-19 | 2007 | 16.20 | kurvits |
| progestin | 20-29 | 2007 | 5.82 | estbb |
| progestin + estrogen | 20-29 | 2007 | 45.08 | estbb |
| progestin | 20-29 | 2007 | 4.40 | kurvits |
| progestin + estrogen | 20-29 | 2007 | 35.30 | kurvits |
| progestin | 30-39 | 2007 | 8.42 | estbb |
| progestin + estrogen | 30-39 | 2007 | 26.83 | estbb |
| progestin | 30-39 | 2007 | 5.60 | kurvits |
| progestin + estrogen | 30-39 | 2007 | 20.10 | kurvits |
| progestin | 40-49 | 2007 | 4.97 | estbb |
| progestin + estrogen | 40-49 | 2007 | 12.81 | estbb |
| progestin | 40-49 | 2007 | 2.90 | kurvits |
| progestin + estrogen | 40-49 | 2007 | 8.10 | kurvits |
| progestin | 50-55 | 2007 | 0.35 | estbb |
| progestin + estrogen | 50-55 | 2007 | 1.76 | estbb |
| progestin | 15-19 | 2008 | 0.46 | estbb |
| progestin + estrogen | 15-19 | 2008 | 22.10 | estbb |
| progestin | 15-19 | 2008 | 0.50 | kurvits |
| progestin + estrogen | 15-19 | 2008 | 16.60 | kurvits |
| progestin | 20-29 | 2008 | 6.47 | estbb |
| progestin + estrogen | 20-29 | 2008 | 44.89 | estbb |
| progestin | 20-29 | 2008 | 5.10 | kurvits |
| progestin + estrogen | 20-29 | 2008 | 35.70 | kurvits |
| progestin | 30-39 | 2008 | 10.65 | estbb |
| progestin + estrogen | 30-39 | 2008 | 27.37 | estbb |
| progestin | 30-39 | 2008 | 7.20 | kurvits |
| progestin + estrogen | 30-39 | 2008 | 20.60 | kurvits |
| progestin | 40-49 | 2008 | 7.05 | estbb |
| progestin + estrogen | 40-49 | 2008 | 12.92 | estbb |
| progestin | 40-49 | 2008 | 4.30 | kurvits |
| progestin + estrogen | 40-49 | 2008 | 8.50 | kurvits |
| progestin | 50-55 | 2008 | 0.82 | estbb |
| progestin + estrogen | 50-55 | 2008 | 1.79 | estbb |
| progestin | 15-19 | 2009 | 0.44 | estbb |
| progestin + estrogen | 15-19 | 2009 | 21.28 | estbb |
| progestin | 15-19 | 2009 | 0.50 | kurvits |
| progestin + estrogen | 15-19 | 2009 | 15.30 | kurvits |
| progestin | 20-29 | 2009 | 6.39 | estbb |
| progestin + estrogen | 20-29 | 2009 | 43.69 | estbb |
| progestin | 20-29 | 2009 | 5.20 | kurvits |
| progestin + estrogen | 20-29 | 2009 | 34.60 | kurvits |
| progestin | 30-39 | 2009 | 11.68 | estbb |
| progestin + estrogen | 30-39 | 2009 | 26.80 | estbb |
| progestin | 30-39 | 2009 | 8.20 | kurvits |
| progestin + estrogen | 30-39 | 2009 | 19.90 | kurvits |
| progestin | 40-49 | 2009 | 8.75 | estbb |
| progestin + estrogen | 40-49 | 2009 | 12.85 | estbb |
| progestin | 40-49 | 2009 | 5.40 | kurvits |
| progestin + estrogen | 40-49 | 2009 | 8.40 | kurvits |
| progestin | 50-55 | 2009 | 1.46 | estbb |
| progestin + estrogen | 50-55 | 2009 | 1.98 | estbb |
| progestin | 15-19 | 2010 | 0.36 | estbb |
| progestin + estrogen | 15-19 | 2010 | 19.98 | estbb |
| progestin | 15-19 | 2010 | 0.40 | kurvits |
| progestin + estrogen | 15-19 | 2010 | 14.20 | kurvits |
| progestin | 20-29 | 2010 | 6.50 | estbb |
| progestin + estrogen | 20-29 | 2010 | 42.10 | estbb |
| progestin | 20-29 | 2010 | 5.20 | kurvits |
| progestin + estrogen | 20-29 | 2010 | 33.20 | kurvits |
| progestin | 30-39 | 2010 | 12.43 | estbb |
| progestin + estrogen | 30-39 | 2010 | 25.15 | estbb |
| progestin | 30-39 | 2010 | 8.80 | kurvits |
| progestin + estrogen | 30-39 | 2010 | 19.30 | kurvits |
| progestin | 40-49 | 2010 | 10.30 | estbb |
| progestin + estrogen | 40-49 | 2010 | 12.33 | estbb |
| progestin | 40-49 | 2010 | 6.30 | kurvits |
| progestin + estrogen | 40-49 | 2010 | 8.40 | kurvits |
| progestin | 50-55 | 2010 | 2.32 | estbb |
| progestin + estrogen | 50-55 | 2010 | 2.35 | estbb |
| progestin | 15-19 | 2011 | 0.41 | estbb |
| progestin + estrogen | 15-19 | 2011 | 21.17 | estbb |
| progestin | 15-19 | 2011 | 0.40 | kurvits |
| progestin + estrogen | 15-19 | 2011 | 15.30 | kurvits |
| progestin | 20-29 | 2011 | 6.36 | estbb |
| progestin + estrogen | 20-29 | 2011 | 43.26 | estbb |
| progestin | 20-29 | 2011 | 4.90 | kurvits |
| progestin + estrogen | 20-29 | 2011 | 34.00 | kurvits |
| progestin | 30-39 | 2011 | 13.24 | estbb |
| progestin + estrogen | 30-39 | 2011 | 25.62 | estbb |
| progestin | 30-39 | 2011 | 9.40 | kurvits |
| progestin + estrogen | 30-39 | 2011 | 20.30 | kurvits |
| progestin | 40-49 | 2011 | 11.44 | estbb |
| progestin + estrogen | 40-49 | 2011 | 12.56 | estbb |
| progestin | 40-49 | 2011 | 7.20 | kurvits |
| progestin + estrogen | 40-49 | 2011 | 8.80 | kurvits |
| progestin | 50-55 | 2011 | 3.23 | estbb |
| progestin + estrogen | 50-55 | 2011 | 2.32 | estbb |
| progestin | 15-19 | 2012 | 0.42 | estbb |
| progestin + estrogen | 15-19 | 2012 | 22.02 | estbb |
| progestin | 15-19 | 2012 | 0.50 | kurvits |
| progestin + estrogen | 15-19 | 2012 | 15.70 | kurvits |
| progestin | 20-29 | 2012 | 5.81 | estbb |
| progestin + estrogen | 20-29 | 2012 | 43.58 | estbb |
| progestin | 20-29 | 2012 | 4.70 | kurvits |
| progestin + estrogen | 20-29 | 2012 | 34.30 | kurvits |
| progestin | 30-39 | 2012 | 13.78 | estbb |
| progestin + estrogen | 30-39 | 2012 | 25.58 | estbb |
| progestin | 30-39 | 2012 | 9.70 | kurvits |
| progestin + estrogen | 30-39 | 2012 | 20.70 | kurvits |
| progestin | 40-49 | 2012 | 12.32 | estbb |
| progestin + estrogen | 40-49 | 2012 | 12.89 | estbb |
| progestin | 40-49 | 2012 | 7.60 | kurvits |
| progestin + estrogen | 40-49 | 2012 | 9.20 | kurvits |
| progestin | 50-55 | 2012 | 3.97 | estbb |
| progestin + estrogen | 50-55 | 2012 | 2.23 | estbb |
| progestin | 15-19 | 2013 | 0.63 | estbb |
| progestin + estrogen | 15-19 | 2013 | 21.68 | estbb |
| progestin | 15-19 | 2013 | 0.60 | kurvits |
| progestin + estrogen | 15-19 | 2013 | 16.10 | kurvits |
| progestin | 20-29 | 2013 | 6.28 | estbb |
| progestin + estrogen | 20-29 | 2013 | 43.24 | estbb |
| progestin | 20-29 | 2013 | 5.00 | kurvits |
| progestin + estrogen | 20-29 | 2013 | 33.10 | kurvits |
| progestin | 30-39 | 2013 | 14.54 | estbb |
| progestin + estrogen | 30-39 | 2013 | 24.37 | estbb |
| progestin | 30-39 | 2013 | 10.50 | kurvits |
| progestin + estrogen | 30-39 | 2013 | 20.20 | kurvits |
| progestin | 40-49 | 2013 | 14.23 | estbb |
| progestin + estrogen | 40-49 | 2013 | 12.43 | estbb |
| progestin | 40-49 | 2013 | 8.80 | kurvits |
| progestin + estrogen | 40-49 | 2013 | 9.10 | kurvits |
| progestin | 50-55 | 2013 | 4.79 | estbb |
| progestin + estrogen | 50-55 | 2013 | 2.17 | estbb |
| progestin | 15-19 | 2014 | 0.84 | estbb |
| progestin + estrogen | 15-19 | 2014 | 21.27 | estbb |
| progestin | 15-19 | 2014 | 0.80 | kurvits |
| progestin + estrogen | 15-19 | 2014 | 16.00 | kurvits |
| progestin | 20-29 | 2014 | 7.14 | estbb |
| progestin + estrogen | 20-29 | 2014 | 41.75 | estbb |
| progestin | 20-29 | 2014 | 5.70 | kurvits |
| progestin + estrogen | 20-29 | 2014 | 31.30 | kurvits |
| progestin | 30-39 | 2014 | 15.88 | estbb |
| progestin + estrogen | 30-39 | 2014 | 22.89 | estbb |
| progestin | 30-39 | 2014 | 11.60 | kurvits |
| progestin + estrogen | 30-39 | 2014 | 19.10 | kurvits |
| progestin | 40-49 | 2014 | 15.93 | estbb |
| progestin + estrogen | 40-49 | 2014 | 11.48 | estbb |
| progestin | 40-49 | 2014 | 10.30 | kurvits |
| progestin + estrogen | 40-49 | 2014 | 8.50 | kurvits |
| progestin | 50-55 | 2014 | 5.68 | estbb |
| progestin + estrogen | 50-55 | 2014 | 2.00 | estbb |
| progestin | 15-19 | 2015 | 1.47 | estbb |
| progestin + estrogen | 15-19 | 2015 | 20.83 | estbb |
| progestin | 15-19 | 2015 | 1.40 | kurvits |
| progestin + estrogen | 15-19 | 2015 | 15.70 | kurvits |
| progestin | 20-29 | 2015 | 8.92 | estbb |
| progestin + estrogen | 20-29 | 2015 | 40.87 | estbb |
| progestin | 20-29 | 2015 | 7.10 | kurvits |
| progestin + estrogen | 20-29 | 2015 | 30.70 | kurvits |
| progestin | 30-39 | 2015 | 17.55 | estbb |
| progestin + estrogen | 30-39 | 2015 | 21.54 | estbb |
| progestin | 30-39 | 2015 | 13.40 | kurvits |
| progestin + estrogen | 30-39 | 2015 | 18.20 | kurvits |
| progestin | 40-49 | 2015 | 18.00 | estbb |
| progestin + estrogen | 40-49 | 2015 | 10.74 | estbb |
| progestin | 40-49 | 2015 | 12.20 | kurvits |
| progestin + estrogen | 40-49 | 2015 | 8.30 | kurvits |
| progestin | 50-55 | 2015 | 6.58 | estbb |
| progestin + estrogen | 50-55 | 2015 | 2.03 | estbb |
| progestin | 15-19 | 2016 | 2.21 | estbb |
| progestin + estrogen | 15-19 | 2016 | 22.63 | estbb |
| progestin | 15-19 | 2016 | 1.70 | kurvits |
| progestin + estrogen | 15-19 | 2016 | 15.20 | kurvits |
| progestin | 20-29 | 2016 | 10.00 | estbb |
| progestin + estrogen | 20-29 | 2016 | 39.92 | estbb |
| progestin | 20-29 | 2016 | 8.10 | kurvits |
| progestin + estrogen | 20-29 | 2016 | 29.30 | kurvits |
| progestin | 30-39 | 2016 | 19.22 | estbb |
| progestin + estrogen | 30-39 | 2016 | 20.57 | estbb |
| progestin | 30-39 | 2016 | 14.90 | kurvits |
| progestin + estrogen | 30-39 | 2016 | 17.60 | kurvits |
| progestin | 40-49 | 2016 | 20.28 | estbb |
| progestin + estrogen | 40-49 | 2016 | 10.28 | estbb |
| progestin | 40-49 | 2016 | 14.00 | kurvits |
| progestin + estrogen | 40-49 | 2016 | 8.40 | kurvits |
| progestin | 50-55 | 2016 | 7.73 | estbb |
| progestin + estrogen | 50-55 | 2016 | 1.75 | estbb |
| progestin | 15-19 | 2017 | 2.93 | estbb |
| progestin + estrogen | 15-19 | 2017 | 25.47 | estbb |
| progestin | 15-19 | 2017 | 1.80 | kurvits |
| progestin + estrogen | 15-19 | 2017 | 15.10 | kurvits |
| progestin | 20-29 | 2017 | 11.21 | estbb |
| progestin + estrogen | 20-29 | 2017 | 37.96 | estbb |
| progestin | 20-29 | 2017 | 8.70 | kurvits |
| progestin + estrogen | 20-29 | 2017 | 27.70 | kurvits |
| progestin | 30-39 | 2017 | 19.73 | estbb |
| progestin + estrogen | 30-39 | 2017 | 20.32 | estbb |
| progestin | 30-39 | 2017 | 15.70 | kurvits |
| progestin + estrogen | 30-39 | 2017 | 17.30 | kurvits |
| progestin | 40-49 | 2017 | 22.00 | estbb |
| progestin + estrogen | 40-49 | 2017 | 10.25 | estbb |
| progestin | 40-49 | 2017 | 15.40 | kurvits |
| progestin + estrogen | 40-49 | 2017 | 8.50 | kurvits |
| progestin | 50-55 | 2017 | 9.69 | estbb |
| progestin + estrogen | 50-55 | 2017 | 1.85 | estbb |
| progestin | 15-19 | 2018 | 3.84 | estbb |
| progestin + estrogen | 15-19 | 2018 | 30.70 | estbb |
| progestin | 15-19 | 2018 | 2.00 | kurvits |
| progestin + estrogen | 15-19 | 2018 | 15.40 | kurvits |
| progestin | 20-29 | 2018 | 12.32 | estbb |
| progestin + estrogen | 20-29 | 2018 | 36.51 | estbb |
| progestin | 20-29 | 2018 | 9.30 | kurvits |
| progestin + estrogen | 20-29 | 2018 | 26.30 | kurvits |
| progestin | 30-39 | 2018 | 20.18 | estbb |
| progestin + estrogen | 30-39 | 2018 | 19.56 | estbb |
| progestin | 30-39 | 2018 | 15.80 | kurvits |
| progestin + estrogen | 30-39 | 2018 | 16.80 | kurvits |
| progestin | 40-49 | 2018 | 23.69 | estbb |
| progestin + estrogen | 40-49 | 2018 | 10.25 | estbb |
| progestin | 40-49 | 2018 | 16.20 | kurvits |
| progestin + estrogen | 40-49 | 2018 | 8.50 | kurvits |
| progestin | 50-55 | 2018 | 11.76 | estbb |
| progestin + estrogen | 50-55 | 2018 | 1.79 | estbb |
| progestin | 15-19 | 2019 | 5.94 | estbb |
| progestin + estrogen | 15-19 | 2019 | 32.35 | estbb |
| progestin | 15-19 | 2019 | 2.50 | kurvits |
| progestin + estrogen | 15-19 | 2019 | 15.20 | kurvits |
| progestin | 20-29 | 2019 | 13.01 | estbb |
| progestin + estrogen | 20-29 | 2019 | 34.58 | estbb |
| progestin | 20-29 | 2019 | 9.90 | kurvits |
| progestin + estrogen | 20-29 | 2019 | 25.50 | kurvits |
| progestin | 30-39 | 2019 | 20.40 | estbb |
| progestin + estrogen | 30-39 | 2019 | 18.74 | estbb |
| progestin | 30-39 | 2019 | 16.20 | kurvits |
| progestin + estrogen | 30-39 | 2019 | 16.30 | kurvits |
| progestin | 40-49 | 2019 | 25.12 | estbb |
| progestin + estrogen | 40-49 | 2019 | 10.16 | estbb |
| progestin | 40-49 | 2019 | 17.30 | kurvits |
| progestin + estrogen | 40-49 | 2019 | 8.50 | kurvits |
| progestin | 50-55 | 2019 | 14.01 | estbb |
| progestin + estrogen | 50-55 | 2019 | 2.03 | estbb |
| progestin | 15-19 | 2020 | 6.72 | estbb |
| progestin + estrogen | 15-19 | 2020 | 33.79 | estbb |
| progestin | 20-29 | 2020 | 14.01 | estbb |
| progestin + estrogen | 20-29 | 2020 | 32.90 | estbb |
| progestin | 30-39 | 2020 | 20.10 | estbb |
| progestin + estrogen | 30-39 | 2020 | 17.25 | estbb |
| progestin | 40-49 | 2020 | 26.11 | estbb |
| progestin + estrogen | 40-49 | 2020 | 9.69 | estbb |
| progestin | 50-55 | 2020 | 15.38 | estbb |
| progestin + estrogen | 50-55 | 2020 | 1.97 | estbb |
| progestin | 15-19 | 2021 | 8.86 | estbb |
| progestin + estrogen | 15-19 | 2021 | 33.54 | estbb |
| progestin | 20-29 | 2021 | 15.10 | estbb |
| progestin + estrogen | 20-29 | 2021 | 29.71 | estbb |
| progestin | 30-39 | 2021 | 20.58 | estbb |
| progestin + estrogen | 30-39 | 2021 | 15.66 | estbb |
| progestin | 40-49 | 2021 | 26.72 | estbb |
| progestin + estrogen | 40-49 | 2021 | 8.82 | estbb |
| progestin | 50-55 | 2021 | 16.57 | estbb |
| progestin + estrogen | 50-55 | 2021 | 1.95 | estbb |
| progestin | 15-19 | 2022 | 10.91 | estbb |
| progestin + estrogen | 15-19 | 2022 | 38.18 | estbb |
| progestin | 20-29 | 2022 | 17.23 | estbb |
| progestin + estrogen | 20-29 | 2022 | 26.56 | estbb |
| progestin | 30-39 | 2022 | 21.28 | estbb |
| progestin + estrogen | 30-39 | 2022 | 14.81 | estbb |
| progestin | 40-49 | 2022 | 27.61 | estbb |
| progestin + estrogen | 40-49 | 2022 | 8.48 | estbb |
| progestin | 50-55 | 2022 | 18.04 | estbb |
| progestin + estrogen | 50-55 | 2022 | 1.94 | estbb |

**Supplementary Table 3.** Proportion of users of different hormonal contraceptive (HC) methods across studied years, stratified by age groups (15–19, 20–29, 30–39, 40–49, 50–55)

| **year** | **hc_type** | **age_group** | **num_users_per_method** | **num_total_uniq_drug_user_combinations** | **proportion** |
| --- | --- | --- | --- | --- | --- |
| 2004 | Anti-androgenic COC | 15-19 | 205 | 2570 | 7.98 |
| 2004 | CHC patch | 15-19 | 121 | 2570 | 4.71 |
| 2004 | CHC ring | 15-19 | 9 | 2570 | 0.35 |
| 2004 | COC 2nd generation | 15-19 | 104 | 2570 | 4.05 |
| 2004 | COC 3rd generation | 15-19 | 1490 | 2570 | 57.98 |
| 2004 | COC 4th generation | 15-19 | 597 | 2570 | 23.23 |
| 2004 | IUD with progestogen | 15-19 | 2 | 2570 | 0.08 |
| 2004 | Progestin-only pill | 15-19 | 42 | 2570 | 1.63 |
| 2005 | Anti-androgenic COC | 15-19 | 203 | 2888 | 7.03 |
| 2005 | CHC patch | 15-19 | 248 | 2888 | 8.59 |
| 2005 | CHC ring | 15-19 | 54 | 2888 | 1.87 |
| 2005 | COC 2nd generation | 15-19 | 164 | 2888 | 5.68 |
| 2005 | COC 3rd generation | 15-19 | 1420 | 2888 | 49.17 |
| 2005 | COC 4th generation | 15-19 | 734 | 2888 | 25.42 |
| 2005 | IUD with progestogen | 15-19 | 1 | 2888 | 0.03 |
| 2005 | Progestin-only pill | 15-19 | 64 | 2888 | 2.22 |
| 2006 | Anti-androgenic COC | 15-19 | 186 | 2943 | 6.32 |
| 2006 | CHC patch | 15-19 | 293 | 2943 | 9.96 |
| 2006 | CHC ring | 15-19 | 103 | 2943 | 3.50 |
| 2006 | COC 2nd generation | 15-19 | 231 | 2943 | 7.85 |
| 2006 | COC 3rd generation | 15-19 | 1233 | 2943 | 41.90 |
| 2006 | COC 4th generation | 15-19 | 845 | 2943 | 28.71 |
| 2006 | IUD with progestogen | 15-19 | 2 | 2943 | 0.07 |
| 2006 | Progestin-only pill | 15-19 | 50 | 2943 | 1.70 |
| 2007 | Anti-androgenic COC | 15-19 | 177 | 3043 | 5.82 |
| 2007 | CHC patch | 15-19 | 326 | 3043 | 10.71 |
| 2007 | CHC ring | 15-19 | 130 | 3043 | 4.27 |
| 2007 | COC 2nd generation | 15-19 | 180 | 3043 | 5.92 |
| 2007 | COC 3rd generation | 15-19 | 947 | 3043 | 31.12 |
| 2007 | COC 4th generation | 15-19 | 1235 | 3043 | 40.58 |
| 2007 | IUD with progestogen | 15-19 | 1 | 3043 | 0.03 |
| 2007 | Progestin-only pill | 15-19 | 47 | 3043 | 1.54 |
| 2008 | Anti-androgenic COC | 15-19 | 149 | 2908 | 5.12 |
| 2008 | CHC patch | 15-19 | 246 | 2908 | 8.46 |
| 2008 | CHC ring | 15-19 | 104 | 2908 | 3.58 |
| 2008 | COC 2nd generation | 15-19 | 167 | 2908 | 5.74 |
| 2008 | COC 3rd generation | 15-19 | 759 | 2908 | 26.10 |
| 2008 | COC 4th generation | 15-19 | 1429 | 2908 | 49.14 |
| 2008 | IUD with progestogen | 15-19 | 3 | 2908 | 0.10 |
| 2008 | Progestin-only pill | 15-19 | 51 | 2908 | 1.75 |
| 2009 | Anti-androgenic COC | 15-19 | 134 | 2562 | 5.23 |
| 2009 | CHC patch | 15-19 | 160 | 2562 | 6.25 |
| 2009 | CHC ring | 15-19 | 103 | 2562 | 4.02 |
| 2009 | COC 2nd generation | 15-19 | 61 | 2562 | 2.38 |
| 2009 | COC 3rd generation | 15-19 | 745 | 2562 | 29.08 |
| 2009 | COC 4th generation | 15-19 | 1313 | 2562 | 51.25 |
| 2009 | IUD with progestogen | 15-19 | 7 | 2562 | 0.27 |
| 2009 | Progestin-only pill | 15-19 | 39 | 2562 | 1.52 |
| 2010 | Anti-androgenic COC | 15-19 | 115 | 2105 | 5.46 |
| 2010 | CHC patch | 15-19 | 120 | 2105 | 5.70 |
| 2010 | CHC ring | 15-19 | 83 | 2105 | 3.94 |
| 2010 | COC 2nd generation | 15-19 | 12 | 2105 | 0.57 |
| 2010 | COC 3rd generation | 15-19 | 717 | 2105 | 34.06 |
| 2010 | COC 4th generation | 15-19 | 1022 | 2105 | 48.55 |
| 2010 | Estradiol COC | 15-19 | 2 | 2105 | 0.10 |
| 2010 | IUD with progestogen | 15-19 | 5 | 2105 | 0.24 |
| 2010 | Progestin-only pill | 15-19 | 29 | 2105 | 1.38 |
| 2011 | Anti-androgenic COC | 15-19 | 95 | 1998 | 4.75 |
| 2011 | CHC patch | 15-19 | 95 | 1998 | 4.75 |
| 2011 | CHC ring | 15-19 | 87 | 1998 | 4.35 |
| 2011 | COC 2nd generation | 15-19 | 6 | 1998 | 0.30 |
| 2011 | COC 3rd generation | 15-19 | 701 | 1998 | 35.09 |
| 2011 | COC 4th generation | 15-19 | 976 | 1998 | 48.85 |
| 2011 | Estradiol COC | 15-19 | 3 | 1998 | 0.15 |
| 2011 | IUD with progestogen | 15-19 | 2 | 1998 | 0.10 |
| 2011 | Progestin-only pill | 15-19 | 33 | 1998 | 1.65 |
| 2012 | Anti-androgenic COC | 15-19 | 106 | 1955 | 5.42 |
| 2012 | CHC patch | 15-19 | 105 | 1955 | 5.37 |
| 2012 | CHC ring | 15-19 | 74 | 1955 | 3.79 |
| 2012 | COC 2nd generation | 15-19 | 3 | 1955 | 0.15 |
| 2012 | COC 3rd generation | 15-19 | 741 | 1955 | 37.90 |
| 2012 | COC 4th generation | 15-19 | 888 | 1955 | 45.42 |
| 2012 | Estradiol COC | 15-19 | 5 | 1955 | 0.26 |
| 2012 | IUD with progestogen | 15-19 | 2 | 1955 | 0.10 |
| 2012 | Progestin-only pill | 15-19 | 31 | 1955 | 1.59 |
| 2013 | Anti-androgenic COC | 15-19 | 107 | 1819 | 5.88 |
| 2013 | CHC patch | 15-19 | 87 | 1819 | 4.78 |
| 2013 | CHC ring | 15-19 | 68 | 1819 | 3.74 |
| 2013 | COC 2nd generation | 15-19 | 3 | 1819 | 0.16 |
| 2013 | COC 3rd generation | 15-19 | 687 | 1819 | 37.77 |
| 2013 | COC 4th generation | 15-19 | 812 | 1819 | 44.64 |
| 2013 | Estradiol COC | 15-19 | 8 | 1819 | 0.44 |
| 2013 | IUD with progestogen | 15-19 | 6 | 1819 | 0.33 |
| 2013 | Progestin-only pill | 15-19 | 41 | 1819 | 2.25 |
| 2014 | Anti-androgenic COC | 15-19 | 94 | 1722 | 5.46 |
| 2014 | CHC patch | 15-19 | 90 | 1722 | 5.23 |
| 2014 | CHC ring | 15-19 | 83 | 1722 | 4.82 |
| 2014 | COC 2nd generation | 15-19 | 7 | 1722 | 0.41 |
| 2014 | COC 3rd generation | 15-19 | 656 | 1722 | 38.10 |
| 2014 | COC 4th generation | 15-19 | 717 | 1722 | 41.64 |
| 2014 | Estradiol COC | 15-19 | 13 | 1722 | 0.75 |
| 2014 | IUD with progestogen | 15-19 | 19 | 1722 | 1.10 |
| 2014 | Implant | 15-19 | 4 | 1722 | 0.23 |
| 2014 | Progestin-only pill | 15-19 | 39 | 1722 | 2.26 |
| 2015 | Anti-androgenic COC | 15-19 | 77 | 1634 | 4.71 |
| 2015 | CHC patch | 15-19 | 94 | 1634 | 5.75 |
| 2015 | CHC ring | 15-19 | 66 | 1634 | 4.04 |
| 2015 | COC 2nd generation | 15-19 | 22 | 1634 | 1.35 |
| 2015 | COC 3rd generation | 15-19 | 648 | 1634 | 39.66 |
| 2015 | COC 4th generation | 15-19 | 617 | 1634 | 37.76 |
| 2015 | Estradiol COC | 15-19 | 11 | 1634 | 0.67 |
| 2015 | IUD with progestogen | 15-19 | 33 | 1634 | 2.02 |
| 2015 | Implant | 15-19 | 19 | 1634 | 1.16 |
| 2015 | Progestin-only pill | 15-19 | 47 | 1634 | 2.88 |
| 2016 | Anti-androgenic COC | 15-19 | 67 | 1530 | 4.38 |
| 2016 | CHC patch | 15-19 | 80 | 1530 | 5.23 |
| 2016 | CHC ring | 15-19 | 74 | 1530 | 4.84 |
| 2016 | COC 2nd generation | 15-19 | 16 | 1530 | 1.05 |
| 2016 | COC 3rd generation | 15-19 | 551 | 1530 | 36.01 |
| 2016 | COC 4th generation | 15-19 | 600 | 1530 | 39.22 |
| 2016 | Estradiol COC | 15-19 | 7 | 1530 | 0.46 |
| 2016 | IUD with progestogen | 15-19 | 56 | 1530 | 3.66 |
| 2016 | Implant | 15-19 | 31 | 1530 | 2.03 |
| 2016 | Progestin-only pill | 15-19 | 48 | 1530 | 3.14 |
| 2017 | Anti-androgenic COC | 15-19 | 50 | 1345 | 3.72 |
| 2017 | CHC patch | 15-19 | 78 | 1345 | 5.80 |
| 2017 | CHC ring | 15-19 | 68 | 1345 | 5.06 |
| 2017 | COC 2nd generation | 15-19 | 12 | 1345 | 0.89 |
| 2017 | COC 3rd generation | 15-19 | 442 | 1345 | 32.86 |
| 2017 | COC 4th generation | 15-19 | 553 | 1345 | 41.12 |
| 2017 | Estradiol COC | 15-19 | 6 | 1345 | 0.45 |
| 2017 | IUD with progestogen | 15-19 | 56 | 1345 | 4.16 |
| 2017 | Implant | 15-19 | 28 | 1345 | 2.08 |
| 2017 | Progestin-only pill | 15-19 | 52 | 1345 | 3.87 |
| 2018 | Anti-androgenic COC | 15-19 | 30 | 1133 | 2.65 |
| 2018 | CHC patch | 15-19 | 68 | 1133 | 6.00 |
| 2018 | CHC ring | 15-19 | 65 | 1133 | 5.74 |
| 2018 | COC 2nd generation | 15-19 | 10 | 1133 | 0.88 |
| 2018 | COC 3rd generation | 15-19 | 322 | 1133 | 28.42 |
| 2018 | COC 4th generation | 15-19 | 510 | 1133 | 45.01 |
| 2018 | Estradiol COC | 15-19 | 5 | 1133 | 0.44 |
| 2018 | IUD with progestogen | 15-19 | 42 | 1133 | 3.71 |
| 2018 | Implant | 15-19 | 31 | 1133 | 2.74 |
| 2018 | Progestin-only pill | 15-19 | 50 | 1133 | 4.41 |
| 2019 | Anti-androgenic COC | 15-19 | 20 | 729 | 2.74 |
| 2019 | CHC patch | 15-19 | 50 | 729 | 6.86 |
| 2019 | CHC ring | 15-19 | 36 | 729 | 4.94 |
| 2019 | COC 2nd generation | 15-19 | 20 | 729 | 2.74 |
| 2019 | COC 3rd generation | 15-19 | 177 | 729 | 24.28 |
| 2019 | COC 4th generation | 15-19 | 314 | 729 | 43.07 |
| 2019 | Estradiol COC | 15-19 | 3 | 729 | 0.41 |
| 2019 | IUD with progestogen | 15-19 | 37 | 729 | 5.08 |
| 2019 | Implant | 15-19 | 33 | 729 | 4.53 |
| 2019 | Progestin-only pill | 15-19 | 39 | 729 | 5.35 |
| 2020 | Anti-androgenic COC | 15-19 | 4 | 257 | 1.56 |
| 2020 | CHC patch | 15-19 | 17 | 257 | 6.61 |
| 2020 | CHC ring | 15-19 | 12 | 257 | 4.67 |
| 2020 | COC 2nd generation | 15-19 | 12 | 257 | 4.67 |
| 2020 | COC 3rd generation | 15-19 | 59 | 257 | 22.96 |
| 2020 | COC 4th generation | 15-19 | 111 | 257 | 43.19 |
| 2020 | Estradiol COC | 15-19 | 2 | 257 | 0.78 |
| 2020 | IUD with progestogen | 15-19 | 16 | 257 | 6.23 |
| 2020 | Implant | 15-19 | 11 | 257 | 4.28 |
| 2020 | Progestin-only pill | 15-19 | 13 | 257 | 5.06 |
| 2021 | Anti-androgenic COC | 15-19 | 2 | 74 | 2.70 |
| 2021 | CHC patch | 15-19 | 4 | 74 | 5.41 |
| 2021 | CHC ring | 15-19 | 6 | 74 | 8.11 |
| 2021 | COC 2nd generation | 15-19 | 5 | 74 | 6.76 |
| 2021 | COC 3rd generation | 15-19 | 11 | 74 | 14.86 |
| 2021 | COC 4th generation | 15-19 | 31 | 74 | 41.89 |
| 2021 | IUD with progestogen | 15-19 | 9 | 74 | 12.16 |
| 2021 | Implant | 15-19 | 1 | 74 | 1.35 |
| 2021 | Progestin-only pill | 15-19 | 5 | 74 | 6.76 |
| 2022 | Anti-androgenic COC | 15-19 | 1 | 29 | 3.45 |
| 2022 | CHC patch | 15-19 | 2 | 29 | 6.90 |
| 2022 | CHC ring | 15-19 | 2 | 29 | 6.90 |
| 2022 | COC 2nd generation | 15-19 | 4 | 29 | 13.79 |
| 2022 | COC 3rd generation | 15-19 | 2 | 29 | 6.90 |
| 2022 | COC 4th generation | 15-19 | 11 | 29 | 37.93 |
| 2022 | IUD with progestogen | 15-19 | 4 | 29 | 13.79 |
| 2022 | Implant | 15-19 | 2 | 29 | 6.90 |
| 2022 | Progestin-only pill | 15-19 | 1 | 29 | 3.45 |
| 2004 | Anti-androgenic COC | 20-29 | 1047 | 12466 | 8.40 |
| 2004 | CHC patch | 20-29 | 788 | 12466 | 6.32 |
| 2004 | CHC ring | 20-29 | 51 | 12466 | 0.41 |
| 2004 | COC 2nd generation | 20-29 | 713 | 12466 | 5.72 |
| 2004 | COC 3rd generation | 20-29 | 6816 | 12466 | 54.68 |
| 2004 | COC 4th generation | 20-29 | 2022 | 12466 | 16.22 |
| 2004 | IUD with progestogen | 20-29 | 74 | 12466 | 0.59 |
| 2004 | Progestin-only pill | 20-29 | 955 | 12466 | 7.66 |
| 2005 | Anti-androgenic COC | 20-29 | 1139 | 15058 | 7.56 |
| 2005 | CHC patch | 20-29 | 1443 | 15058 | 9.58 |
| 2005 | CHC ring | 20-29 | 548 | 15058 | 3.64 |
| 2005 | COC 2nd generation | 20-29 | 773 | 15058 | 5.13 |
| 2005 | COC 3rd generation | 20-29 | 6772 | 15058 | 44.97 |
| 2005 | COC 4th generation | 20-29 | 2955 | 15058 | 19.62 |
| 2005 | IUD with progestogen | 20-29 | 180 | 15058 | 1.20 |
| 2005 | Progestin-only pill | 20-29 | 1248 | 15058 | 8.29 |
| 2006 | Anti-androgenic COC | 20-29 | 928 | 15158 | 6.12 |
| 2006 | CHC patch | 20-29 | 1586 | 15158 | 10.46 |
| 2006 | CHC ring | 20-29 | 1011 | 15158 | 6.67 |
| 2006 | COC 2nd generation | 20-29 | 806 | 15158 | 5.32 |
| 2006 | COC 3rd generation | 20-29 | 6091 | 15158 | 40.18 |
| 2006 | COC 4th generation | 20-29 | 3223 | 15158 | 21.26 |
| 2006 | IUD with progestogen | 20-29 | 299 | 15158 | 1.97 |
| 2006 | Progestin-only pill | 20-29 | 1214 | 15158 | 8.01 |
| 2007 | Anti-androgenic COC | 20-29 | 834 | 15129 | 5.51 |
| 2007 | CHC patch | 20-29 | 1595 | 15129 | 10.54 |
| 2007 | CHC ring | 20-29 | 1141 | 15129 | 7.54 |
| 2007 | COC 2nd generation | 20-29 | 616 | 15129 | 4.07 |
| 2007 | COC 3rd generation | 20-29 | 5255 | 15129 | 34.73 |
| 2007 | COC 4th generation | 20-29 | 4135 | 15129 | 27.33 |
| 2007 | IUD with progestogen | 20-29 | 370 | 15129 | 2.45 |
| 2007 | Progestin-only pill | 20-29 | 1183 | 15129 | 7.82 |
| 2008 | Anti-androgenic COC | 20-29 | 822 | 15144 | 5.43 |
| 2008 | CHC patch | 20-29 | 1453 | 15144 | 9.59 |
| 2008 | CHC ring | 20-29 | 1249 | 15144 | 8.25 |
| 2008 | COC 2nd generation | 20-29 | 492 | 15144 | 3.25 |
| 2008 | COC 3rd generation | 20-29 | 4562 | 15144 | 30.12 |
| 2008 | COC 4th generation | 20-29 | 4799 | 15144 | 31.69 |
| 2008 | IUD with progestogen | 20-29 | 460 | 15144 | 3.04 |
| 2008 | Progestin-only pill | 20-29 | 1307 | 15144 | 8.63 |
| 2009 | Anti-androgenic COC | 20-29 | 754 | 14882 | 5.07 |
| 2009 | CHC patch | 20-29 | 1175 | 14882 | 7.90 |
| 2009 | CHC ring | 20-29 | 1316 | 14882 | 8.84 |
| 2009 | COC 2nd generation | 20-29 | 354 | 14882 | 2.38 |
| 2009 | COC 3rd generation | 20-29 | 4205 | 14882 | 28.26 |
| 2009 | COC 4th generation | 20-29 | 5331 | 14882 | 35.82 |
| 2009 | IUD with progestogen | 20-29 | 505 | 14882 | 3.39 |
| 2009 | Progestin-only pill | 20-29 | 1242 | 14882 | 8.35 |
| 2010 | Anti-androgenic COC | 20-29 | 688 | 14316 | 4.81 |
| 2010 | CHC patch | 20-29 | 993 | 14316 | 6.94 |
| 2010 | CHC ring | 20-29 | 1301 | 14316 | 9.09 |
| 2010 | COC 2nd generation | 20-29 | 64 | 14316 | 0.45 |
| 2010 | COC 3rd generation | 20-29 | 4031 | 14316 | 28.16 |
| 2010 | COC 4th generation | 20-29 | 5431 | 14316 | 37.94 |
| 2010 | Estradiol COC | 20-29 | 17 | 14316 | 0.12 |
| 2010 | IUD with progestogen | 20-29 | 525 | 14316 | 3.67 |
| 2010 | Progestin-only pill | 20-29 | 1266 | 14316 | 8.84 |
| 2011 | Anti-androgenic COC | 20-29 | 658 | 14310 | 4.60 |
| 2011 | CHC patch | 20-29 | 908 | 14310 | 6.35 |
| 2011 | CHC ring | 20-29 | 1347 | 14310 | 9.41 |
| 2011 | COC 2nd generation | 20-29 | 44 | 14310 | 0.31 |
| 2011 | COC 3rd generation | 20-29 | 3923 | 14310 | 27.41 |
| 2011 | COC 4th generation | 20-29 | 5668 | 14310 | 39.61 |
| 2011 | Estradiol COC | 20-29 | 22 | 14310 | 0.15 |
| 2011 | IUD with progestogen | 20-29 | 569 | 14310 | 3.98 |
| 2011 | Progestin-only pill | 20-29 | 1171 | 14310 | 8.18 |
| 2012 | Anti-androgenic COC | 20-29 | 735 | 14057 | 5.23 |
| 2012 | CHC patch | 20-29 | 850 | 14057 | 6.05 |
| 2012 | CHC ring | 20-29 | 1282 | 14057 | 9.12 |
| 2012 | COC 2nd generation | 20-29 | 43 | 14057 | 0.31 |
| 2012 | COC 3rd generation | 20-29 | 3995 | 14057 | 28.42 |
| 2012 | COC 4th generation | 20-29 | 5545 | 14057 | 39.45 |
| 2012 | Estradiol COC | 20-29 | 56 | 14057 | 0.40 |
| 2012 | IUD with progestogen | 20-29 | 524 | 14057 | 3.73 |
| 2012 | Progestin-only pill | 20-29 | 1027 | 14057 | 7.31 |
| 2013 | Anti-androgenic COC | 20-29 | 762 | 13419 | 5.68 |
| 2013 | CHC patch | 20-29 | 801 | 13419 | 5.97 |
| 2013 | CHC ring | 20-29 | 1146 | 13419 | 8.54 |
| 2013 | COC 2nd generation | 20-29 | 35 | 13419 | 0.26 |
| 2013 | COC 3rd generation | 20-29 | 3808 | 13419 | 28.38 |
| 2013 | COC 4th generation | 20-29 | 5160 | 13419 | 38.45 |
| 2013 | Estradiol COC | 20-29 | 80 | 13419 | 0.60 |
| 2013 | IUD with progestogen | 20-29 | 638 | 13419 | 4.75 |
| 2013 | Progestin-only pill | 20-29 | 989 | 13419 | 7.37 |
| 2014 | Anti-androgenic COC | 20-29 | 653 | 12575 | 5.19 |
| 2014 | CHC patch | 20-29 | 707 | 12575 | 5.62 |
| 2014 | CHC ring | 20-29 | 1030 | 12575 | 8.19 |
| 2014 | COC 2nd generation | 20-29 | 50 | 12575 | 0.40 |
| 2014 | COC 3rd generation | 20-29 | 3546 | 12575 | 28.20 |
| 2014 | COC 4th generation | 20-29 | 4761 | 12575 | 37.86 |
| 2014 | Estradiol COC | 20-29 | 52 | 12575 | 0.41 |
| 2014 | IUD with progestogen | 20-29 | 816 | 12575 | 6.49 |
| 2014 | Implant | 20-29 | 25 | 12575 | 0.20 |
| 2014 | Progestin-only pill | 20-29 | 935 | 12575 | 7.44 |
| 2015 | Anti-androgenic COC | 20-29 | 607 | 12269 | 4.95 |
| 2015 | CHC patch | 20-29 | 624 | 12269 | 5.09 |
| 2015 | CHC ring | 20-29 | 944 | 12269 | 7.69 |
| 2015 | COC 2nd generation | 20-29 | 104 | 12269 | 0.85 |
| 2015 | COC 3rd generation | 20-29 | 3343 | 12269 | 27.25 |
| 2015 | COC 4th generation | 20-29 | 4467 | 12269 | 36.41 |
| 2015 | Estradiol COC | 20-29 | 46 | 12269 | 0.37 |
| 2015 | IUD with progestogen | 20-29 | 1025 | 12269 | 8.35 |
| 2015 | Implant | 20-29 | 146 | 12269 | 1.19 |
| 2015 | Progestin-only pill | 20-29 | 963 | 12269 | 7.85 |
| 2016 | Anti-androgenic COC | 20-29 | 480 | 11613 | 4.13 |
| 2016 | CHC patch | 20-29 | 654 | 11613 | 5.63 |
| 2016 | CHC ring | 20-29 | 860 | 11613 | 7.41 |
| 2016 | COC 2nd generation | 20-29 | 95 | 11613 | 0.82 |
| 2016 | COC 3rd generation | 20-29 | 3052 | 11613 | 26.28 |
| 2016 | COC 4th generation | 20-29 | 4161 | 11613 | 35.83 |
| 2016 | Estradiol COC | 20-29 | 40 | 11613 | 0.34 |
| 2016 | IUD with progestogen | 20-29 | 1147 | 11613 | 9.88 |
| 2016 | Implant | 20-29 | 245 | 11613 | 2.11 |
| 2016 | Progestin-only pill | 20-29 | 879 | 11613 | 7.57 |
| 2017 | Anti-androgenic COC | 20-29 | 435 | 10681 | 4.07 |
| 2017 | CHC patch | 20-29 | 535 | 10681 | 5.01 |
| 2017 | CHC ring | 20-29 | 812 | 10681 | 7.60 |
| 2017 | COC 2nd generation | 20-29 | 105 | 10681 | 0.98 |
| 2017 | COC 3rd generation | 20-29 | 2678 | 10681 | 25.07 |
| 2017 | COC 4th generation | 20-29 | 3679 | 10681 | 34.44 |
| 2017 | Estradiol COC | 20-29 | 46 | 10681 | 0.43 |
| 2017 | IUD with progestogen | 20-29 | 1293 | 10681 | 12.11 |
| 2017 | Implant | 20-29 | 302 | 10681 | 2.83 |
| 2017 | Progestin-only pill | 20-29 | 796 | 10681 | 7.45 |
| 2018 | Anti-androgenic COC | 20-29 | 391 | 9926 | 3.94 |
| 2018 | CHC patch | 20-29 | 490 | 9926 | 4.94 |
| 2018 | CHC ring | 20-29 | 734 | 9926 | 7.39 |
| 2018 | COC 2nd generation | 20-29 | 124 | 9926 | 1.25 |
| 2018 | COC 3rd generation | 20-29 | 2300 | 9926 | 23.17 |
| 2018 | COC 4th generation | 20-29 | 3406 | 9926 | 34.31 |
| 2018 | Estradiol COC | 20-29 | 35 | 9926 | 0.35 |
| 2018 | IUD with progestogen | 20-29 | 1239 | 9926 | 12.48 |
| 2018 | Implant | 20-29 | 366 | 9926 | 3.69 |
| 2018 | Progestin-only pill | 20-29 | 841 | 9926 | 8.47 |
| 2019 | Anti-androgenic COC | 20-29 | 324 | 8825 | 3.67 |
| 2019 | CHC patch | 20-29 | 443 | 8825 | 5.02 |
| 2019 | CHC ring | 20-29 | 644 | 8825 | 7.30 |
| 2019 | COC 2nd generation | 20-29 | 132 | 8825 | 1.50 |
| 2019 | COC 3rd generation | 20-29 | 1916 | 8825 | 21.71 |
| 2019 | COC 4th generation | 20-29 | 2974 | 8825 | 33.70 |
| 2019 | Estradiol COC | 20-29 | 25 | 8825 | 0.28 |
| 2019 | IUD with progestogen | 20-29 | 1223 | 8825 | 13.86 |
| 2019 | Implant | 20-29 | 383 | 8825 | 4.34 |
| 2019 | Progestin-only pill | 20-29 | 761 | 8825 | 8.62 |
| 2020 | Anti-androgenic COC | 20-29 | 276 | 7974 | 3.46 |
| 2020 | CHC patch | 20-29 | 406 | 7974 | 5.09 |
| 2020 | CHC ring | 20-29 | 545 | 7974 | 6.83 |
| 2020 | COC 2nd generation | 20-29 | 157 | 7974 | 1.97 |
| 2020 | COC 3rd generation | 20-29 | 1630 | 7974 | 20.44 |
| 2020 | COC 4th generation | 20-29 | 2592 | 7974 | 32.51 |
| 2020 | Estradiol COC | 20-29 | 23 | 7974 | 0.29 |
| 2020 | IUD with progestogen | 20-29 | 1240 | 7974 | 15.55 |
| 2020 | Implant | 20-29 | 421 | 7974 | 5.28 |
| 2020 | Progestin-only pill | 20-29 | 684 | 7974 | 8.58 |
| 2021 | Anti-androgenic COC | 20-29 | 209 | 6728 | 3.11 |
| 2021 | CHC patch | 20-29 | 346 | 6728 | 5.14 |
| 2021 | CHC ring | 20-29 | 470 | 6728 | 6.99 |
| 2021 | COC 2nd generation | 20-29 | 157 | 6728 | 2.33 |
| 2021 | COC 3rd generation | 20-29 | 1250 | 6728 | 18.58 |
| 2021 | COC 4th generation | 20-29 | 2057 | 6728 | 30.57 |
| 2021 | Estradiol COC | 20-29 | 19 | 6728 | 0.28 |
| 2021 | IUD with progestogen | 20-29 | 1223 | 6728 | 18.18 |
| 2021 | Implant | 20-29 | 403 | 6728 | 5.99 |
| 2021 | Progestin-only pill | 20-29 | 594 | 6728 | 8.83 |
| 2022 | Anti-androgenic COC | 20-29 | 165 | 5558 | 2.97 |
| 2022 | CHC patch | 20-29 | 243 | 5558 | 4.37 |
| 2022 | CHC ring | 20-29 | 349 | 5558 | 6.28 |
| 2022 | COC 2nd generation | 20-29 | 129 | 5558 | 2.32 |
| 2022 | COC 3rd generation | 20-29 | 901 | 5558 | 16.21 |
| 2022 | COC 4th generation | 20-29 | 1582 | 5558 | 28.46 |
| 2022 | Estradiol COC | 20-29 | 33 | 5558 | 0.59 |
| 2022 | IUD with progestogen | 20-29 | 1237 | 5558 | 22.26 |
| 2022 | Implant | 20-29 | 354 | 5558 | 6.37 |
| 2022 | Progestin-only pill | 20-29 | 565 | 5558 | 10.17 |
| 2004 | Anti-androgenic COC | 30-39 | 501 | 6890 | 7.27 |
| 2004 | CHC patch | 30-39 | 637 | 6890 | 9.25 |
| 2004 | CHC ring | 30-39 | 36 | 6890 | 0.52 |
| 2004 | COC 2nd generation | 30-39 | 684 | 6890 | 9.93 |
| 2004 | COC 3rd generation | 30-39 | 3127 | 6890 | 45.38 |
| 2004 | COC 4th generation | 30-39 | 1070 | 6890 | 15.53 |
| 2004 | IUD with progestogen | 30-39 | 264 | 6890 | 3.83 |
| 2004 | Progestin-only pill | 30-39 | 571 | 6890 | 8.29 |
| 2005 | Anti-androgenic COC | 30-39 | 492 | 8877 | 5.54 |
| 2005 | CHC patch | 30-39 | 1059 | 8877 | 11.93 |
| 2005 | CHC ring | 30-39 | 483 | 8877 | 5.44 |
| 2005 | COC 2nd generation | 30-39 | 722 | 8877 | 8.13 |
| 2005 | COC 3rd generation | 30-39 | 3226 | 8877 | 36.34 |
| 2005 | COC 4th generation | 30-39 | 1424 | 8877 | 16.04 |
| 2005 | IUD with progestogen | 30-39 | 602 | 8877 | 6.78 |
| 2005 | Progestin-only pill | 30-39 | 869 | 8877 | 9.79 |
| 2006 | Anti-androgenic COC | 30-39 | 460 | 9535 | 4.82 |
| 2006 | CHC patch | 30-39 | 1057 | 9535 | 11.09 |
| 2006 | CHC ring | 30-39 | 821 | 9535 | 8.61 |
| 2006 | COC 2nd generation | 30-39 | 679 | 9535 | 7.12 |
| 2006 | COC 3rd generation | 30-39 | 3017 | 9535 | 31.64 |
| 2006 | COC 4th generation | 30-39 | 1614 | 9535 | 16.93 |
| 2006 | IUD with progestogen | 30-39 | 968 | 9535 | 10.15 |
| 2006 | Progestin-only pill | 30-39 | 919 | 9535 | 9.64 |
| 2007 | Anti-androgenic COC | 30-39 | 465 | 10121 | 4.59 |
| 2007 | CHC patch | 30-39 | 1031 | 10121 | 10.19 |
| 2007 | CHC ring | 30-39 | 997 | 10121 | 9.85 |
| 2007 | COC 2nd generation | 30-39 | 549 | 10121 | 5.42 |
| 2007 | COC 3rd generation | 30-39 | 2861 | 10121 | 28.27 |
| 2007 | COC 4th generation | 30-39 | 1948 | 10121 | 19.25 |
| 2007 | IUD with progestogen | 30-39 | 1367 | 10121 | 13.51 |
| 2007 | Progestin-only pill | 30-39 | 903 | 10121 | 8.92 |
| 2008 | Anti-androgenic COC | 30-39 | 459 | 10785 | 4.26 |
| 2008 | CHC patch | 30-39 | 984 | 10785 | 9.12 |
| 2008 | CHC ring | 30-39 | 1135 | 10785 | 10.52 |
| 2008 | COC 2nd generation | 30-39 | 468 | 10785 | 4.34 |
| 2008 | COC 3rd generation | 30-39 | 2654 | 10785 | 24.61 |
| 2008 | COC 4th generation | 30-39 | 2208 | 10785 | 20.47 |
| 2008 | IUD with progestogen | 30-39 | 1813 | 10785 | 16.81 |
| 2008 | Progestin-only pill | 30-39 | 1064 | 10785 | 9.87 |
| 2009 | Anti-androgenic COC | 30-39 | 406 | 10785 | 3.76 |
| 2009 | CHC patch | 30-39 | 830 | 10785 | 7.70 |
| 2009 | CHC ring | 30-39 | 1155 | 10785 | 10.71 |
| 2009 | COC 2nd generation | 30-39 | 364 | 10785 | 3.38 |
| 2009 | COC 3rd generation | 30-39 | 2502 | 10785 | 23.20 |
| 2009 | COC 4th generation | 30-39 | 2398 | 10785 | 22.23 |
| 2009 | IUD with progestogen | 30-39 | 2064 | 10785 | 19.14 |
| 2009 | Progestin-only pill | 30-39 | 1066 | 10785 | 9.88 |
| 2010 | Anti-androgenic COC | 30-39 | 357 | 10383 | 3.44 |
| 2010 | CHC patch | 30-39 | 660 | 10383 | 6.36 |
| 2010 | CHC ring | 30-39 | 1163 | 10383 | 11.20 |
| 2010 | COC 2nd generation | 30-39 | 234 | 10383 | 2.25 |
| 2010 | COC 3rd generation | 30-39 | 2301 | 10383 | 22.16 |
| 2010 | COC 4th generation | 30-39 | 2333 | 10383 | 22.47 |
| 2010 | Estradiol COC | 30-39 | 24 | 10383 | 0.23 |
| 2010 | IUD with progestogen | 30-39 | 2214 | 10383 | 21.32 |
| 2010 | Progestin-only pill | 30-39 | 1097 | 10383 | 10.57 |
| 2011 | Anti-androgenic COC | 30-39 | 323 | 10670 | 3.03 |
| 2011 | CHC patch | 30-39 | 637 | 10670 | 5.97 |
| 2011 | CHC ring | 30-39 | 1199 | 10670 | 11.24 |
| 2011 | COC 2nd generation | 30-39 | 191 | 10670 | 1.79 |
| 2011 | COC 3rd generation | 30-39 | 2305 | 10670 | 21.60 |
| 2011 | COC 4th generation | 30-39 | 2467 | 10670 | 23.12 |
| 2011 | Estradiol COC | 30-39 | 29 | 10670 | 0.27 |
| 2011 | IUD with progestogen | 30-39 | 2370 | 10670 | 22.21 |
| 2011 | Progestin-only pill | 30-39 | 1149 | 10670 | 10.77 |
| 2012 | Anti-androgenic COC | 30-39 | 328 | 10713 | 3.06 |
| 2012 | CHC patch | 30-39 | 612 | 10713 | 5.71 |
| 2012 | CHC ring | 30-39 | 1174 | 10713 | 10.96 |
| 2012 | COC 2nd generation | 30-39 | 160 | 10713 | 1.49 |
| 2012 | COC 3rd generation | 30-39 | 2263 | 10713 | 21.12 |
| 2012 | COC 4th generation | 30-39 | 2464 | 10713 | 23.00 |
| 2012 | Estradiol COC | 30-39 | 61 | 10713 | 0.57 |
| 2012 | IUD with progestogen | 30-39 | 2498 | 10713 | 23.32 |
| 2012 | Progestin-only pill | 30-39 | 1153 | 10713 | 10.76 |
| 2013 | Anti-androgenic COC | 30-39 | 360 | 10669 | 3.37 |
| 2013 | CHC patch | 30-39 | 581 | 10669 | 5.45 |
| 2013 | CHC ring | 30-39 | 1043 | 10669 | 9.78 |
| 2013 | COC 2nd generation | 30-39 | 126 | 10669 | 1.18 |
| 2013 | COC 3rd generation | 30-39 | 2171 | 10669 | 20.35 |
| 2013 | COC 4th generation | 30-39 | 2404 | 10669 | 22.53 |
| 2013 | Estradiol COC | 30-39 | 83 | 10669 | 0.78 |
| 2013 | IUD with progestogen | 30-39 | 2888 | 10669 | 27.07 |
| 2013 | Progestin-only pill | 30-39 | 1013 | 10669 | 9.49 |
| 2014 | Anti-androgenic COC | 30-39 | 345 | 10605 | 3.25 |
| 2014 | CHC patch | 30-39 | 512 | 10605 | 4.83 |
| 2014 | CHC ring | 30-39 | 972 | 10605 | 9.17 |
| 2014 | COC 2nd generation | 30-39 | 100 | 10605 | 0.94 |
| 2014 | COC 3rd generation | 30-39 | 2093 | 10605 | 19.74 |
| 2014 | COC 4th generation | 30-39 | 2257 | 10605 | 21.28 |
| 2014 | Estradiol COC | 30-39 | 72 | 10605 | 0.68 |
| 2014 | IUD with progestogen | 30-39 | 3277 | 10605 | 30.90 |
| 2014 | Implant | 30-39 | 17 | 10605 | 0.16 |
| 2014 | Progestin-only pill | 30-39 | 960 | 10605 | 9.05 |
| 2015 | Anti-androgenic COC | 30-39 | 277 | 10712 | 2.59 |
| 2015 | CHC patch | 30-39 | 492 | 10712 | 4.59 |
| 2015 | CHC ring | 30-39 | 897 | 10712 | 8.37 |
| 2015 | COC 2nd generation | 30-39 | 85 | 10712 | 0.79 |
| 2015 | COC 3rd generation | 30-39 | 1951 | 10712 | 18.21 |
| 2015 | COC 4th generation | 30-39 | 2211 | 10712 | 20.64 |
| 2015 | Estradiol COC | 30-39 | 55 | 10712 | 0.51 |
| 2015 | IUD with progestogen | 30-39 | 3667 | 10712 | 34.23 |
| 2015 | Implant | 30-39 | 111 | 10712 | 1.04 |
| 2015 | Progestin-only pill | 30-39 | 966 | 10712 | 9.02 |
| 2016 | Anti-androgenic COC | 30-39 | 259 | 10908 | 2.37 |
| 2016 | CHC patch | 30-39 | 472 | 10908 | 4.33 |
| 2016 | CHC ring | 30-39 | 861 | 10908 | 7.89 |
| 2016 | COC 2nd generation | 30-39 | 73 | 10908 | 0.67 |
| 2016 | COC 3rd generation | 30-39 | 1857 | 10908 | 17.02 |
| 2016 | COC 4th generation | 30-39 | 2154 | 10908 | 19.75 |
| 2016 | Estradiol COC | 30-39 | 47 | 10908 | 0.43 |
| 2016 | IUD with progestogen | 30-39 | 4012 | 10908 | 36.78 |
| 2016 | Implant | 30-39 | 196 | 10908 | 1.80 |
| 2016 | Progestin-only pill | 30-39 | 977 | 10908 | 8.96 |
| 2017 | Anti-androgenic COC | 30-39 | 233 | 10952 | 2.13 |
| 2017 | CHC patch | 30-39 | 416 | 10952 | 3.80 |
| 2017 | CHC ring | 30-39 | 831 | 10952 | 7.59 |
| 2017 | COC 2nd generation | 30-39 | 79 | 10952 | 0.72 |
| 2017 | COC 3rd generation | 30-39 | 1841 | 10952 | 16.81 |
| 2017 | COC 4th generation | 30-39 | 2193 | 10952 | 20.02 |
| 2017 | Estradiol COC | 30-39 | 44 | 10952 | 0.40 |
| 2017 | IUD with progestogen | 30-39 | 4146 | 10952 | 37.86 |
| 2017 | Implant | 30-39 | 209 | 10952 | 1.91 |
| 2017 | Progestin-only pill | 30-39 | 960 | 10952 | 8.77 |
| 2018 | Anti-androgenic COC | 30-39 | 190 | 10934 | 1.74 |
| 2018 | CHC patch | 30-39 | 392 | 10934 | 3.59 |
| 2018 | CHC ring | 30-39 | 803 | 10934 | 7.34 |
| 2018 | COC 2nd generation | 30-39 | 84 | 10934 | 0.77 |
| 2018 | COC 3rd generation | 30-39 | 1729 | 10934 | 15.81 |
| 2018 | COC 4th generation | 30-39 | 2197 | 10934 | 20.09 |
| 2018 | Estradiol COC | 30-39 | 41 | 10934 | 0.37 |
| 2018 | IUD with progestogen | 30-39 | 4228 | 10934 | 38.67 |
| 2018 | Implant | 30-39 | 224 | 10934 | 2.05 |
| 2018 | Progestin-only pill | 30-39 | 1046 | 10934 | 9.57 |
| 2019 | Anti-androgenic COC | 30-39 | 173 | 10871 | 1.59 |
| 2019 | CHC patch | 30-39 | 398 | 10871 | 3.66 |
| 2019 | CHC ring | 30-39 | 768 | 10871 | 7.06 |
| 2019 | COC 2nd generation | 30-39 | 88 | 10871 | 0.81 |
| 2019 | COC 3rd generation | 30-39 | 1650 | 10871 | 15.18 |
| 2019 | COC 4th generation | 30-39 | 2152 | 10871 | 19.80 |
| 2019 | Estradiol COC | 30-39 | 33 | 10871 | 0.30 |
| 2019 | IUD with progestogen | 30-39 | 4297 | 10871 | 39.53 |
| 2019 | Implant | 30-39 | 229 | 10871 | 2.11 |
| 2019 | Progestin-only pill | 30-39 | 1083 | 10871 | 9.96 |
| 2020 | Anti-androgenic COC | 30-39 | 157 | 10369 | 1.51 |
| 2020 | CHC patch | 30-39 | 373 | 10369 | 3.60 |
| 2020 | CHC ring | 30-39 | 682 | 10369 | 6.58 |
| 2020 | COC 2nd generation | 30-39 | 62 | 10369 | 0.60 |
| 2020 | COC 3rd generation | 30-39 | 1468 | 10369 | 14.16 |
| 2020 | COC 4th generation | 30-39 | 2060 | 10369 | 19.87 |
| 2020 | Estradiol COC | 30-39 | 30 | 10369 | 0.29 |
| 2020 | IUD with progestogen | 30-39 | 4215 | 10369 | 40.65 |
| 2020 | Implant | 30-39 | 225 | 10369 | 2.17 |
| 2020 | Progestin-only pill | 30-39 | 1097 | 10369 | 10.58 |
| 2021 | Anti-androgenic COC | 30-39 | 144 | 9954 | 1.45 |
| 2021 | CHC patch | 30-39 | 333 | 9954 | 3.35 |
| 2021 | CHC ring | 30-39 | 594 | 9954 | 5.97 |
| 2021 | COC 2nd generation | 30-39 | 60 | 9954 | 0.60 |
| 2021 | COC 3rd generation | 30-39 | 1316 | 9954 | 13.22 |
| 2021 | COC 4th generation | 30-39 | 1863 | 9954 | 18.72 |
| 2021 | Estradiol COC | 30-39 | 28 | 9954 | 0.28 |
| 2021 | IUD with progestogen | 30-39 | 4279 | 9954 | 42.99 |
| 2021 | Implant | 30-39 | 227 | 9954 | 2.28 |
| 2021 | Progestin-only pill | 30-39 | 1110 | 9954 | 11.15 |
| 2022 | Anti-androgenic COC | 30-39 | 129 | 9711 | 1.33 |
| 2022 | CHC patch | 30-39 | 317 | 9711 | 3.26 |
| 2022 | CHC ring | 30-39 | 550 | 9711 | 5.66 |
| 2022 | COC 2nd generation | 30-39 | 59 | 9711 | 0.61 |
| 2022 | COC 3rd generation | 30-39 | 1160 | 9711 | 11.95 |
| 2022 | COC 4th generation | 30-39 | 1742 | 9711 | 17.94 |
| 2022 | Estradiol COC | 30-39 | 47 | 9711 | 0.48 |
| 2022 | IUD with progestogen | 30-39 | 4297 | 9711 | 44.25 |
| 2022 | Implant | 30-39 | 222 | 9711 | 2.29 |
| 2022 | Progestin-only pill | 30-39 | 1188 | 9711 | 12.23 |
| 2004 | Anti-androgenic COC | 40-49 | 141 | 2482 | 5.68 |
| 2004 | CHC patch | 40-49 | 179 | 2482 | 7.21 |
| 2004 | CHC ring | 40-49 | 7 | 2482 | 0.28 |
| 2004 | COC 2nd generation | 40-49 | 323 | 2482 | 13.01 |
| 2004 | COC 3rd generation | 40-49 | 1345 | 2482 | 54.19 |
| 2004 | COC 4th generation | 40-49 | 288 | 2482 | 11.60 |
| 2004 | IUD with progestogen | 40-49 | 129 | 2482 | 5.20 |
| 2004 | Progestin-only pill | 40-49 | 70 | 2482 | 2.82 |
| 2005 | Anti-androgenic COC | 40-49 | 158 | 3271 | 4.83 |
| 2005 | CHC patch | 40-49 | 305 | 3271 | 9.32 |
| 2005 | CHC ring | 40-49 | 143 | 3271 | 4.37 |
| 2005 | COC 2nd generation | 40-49 | 347 | 3271 | 10.61 |
| 2005 | COC 3rd generation | 40-49 | 1421 | 3271 | 43.44 |
| 2005 | COC 4th generation | 40-49 | 440 | 3271 | 13.45 |
| 2005 | IUD with progestogen | 40-49 | 358 | 3271 | 10.94 |
| 2005 | Progestin-only pill | 40-49 | 99 | 3271 | 3.03 |
| 2006 | Anti-androgenic COC | 40-49 | 145 | 3758 | 3.86 |
| 2006 | CHC patch | 40-49 | 333 | 3758 | 8.86 |
| 2006 | CHC ring | 40-49 | 295 | 3758 | 7.85 |
| 2006 | COC 2nd generation | 40-49 | 341 | 3758 | 9.07 |
| 2006 | COC 3rd generation | 40-49 | 1402 | 3758 | 37.31 |
| 2006 | COC 4th generation | 40-49 | 513 | 3758 | 13.65 |
| 2006 | IUD with progestogen | 40-49 | 630 | 3758 | 16.76 |
| 2006 | Progestin-only pill | 40-49 | 99 | 3758 | 2.63 |
| 2007 | Anti-androgenic COC | 40-49 | 156 | 4396 | 3.55 |
| 2007 | CHC patch | 40-49 | 365 | 4396 | 8.30 |
| 2007 | CHC ring | 40-49 | 395 | 4396 | 8.99 |
| 2007 | COC 2nd generation | 40-49 | 305 | 4396 | 6.94 |
| 2007 | COC 3rd generation | 40-49 | 1336 | 4396 | 30.39 |
| 2007 | COC 4th generation | 40-49 | 684 | 4396 | 15.56 |
| 2007 | IUD with progestogen | 40-49 | 1035 | 4396 | 23.54 |
| 2007 | Progestin-only pill | 40-49 | 120 | 4396 | 2.73 |
| 2008 | Anti-androgenic COC | 40-49 | 160 | 4918 | 3.25 |
| 2008 | CHC patch | 40-49 | 361 | 4918 | 7.34 |
| 2008 | CHC ring | 40-49 | 434 | 4918 | 8.82 |
| 2008 | COC 2nd generation | 40-49 | 270 | 4918 | 5.49 |
| 2008 | COC 3rd generation | 40-49 | 1221 | 4918 | 24.83 |
| 2008 | COC 4th generation | 40-49 | 802 | 4918 | 16.31 |
| 2008 | IUD with progestogen | 40-49 | 1515 | 4918 | 30.81 |
| 2008 | Progestin-only pill | 40-49 | 155 | 4918 | 3.15 |
| 2009 | Anti-androgenic COC | 40-49 | 152 | 5377 | 2.83 |
| 2009 | CHC patch | 40-49 | 322 | 5377 | 5.99 |
| 2009 | CHC ring | 40-49 | 469 | 5377 | 8.72 |
| 2009 | COC 2nd generation | 40-49 | 247 | 5377 | 4.59 |
| 2009 | COC 3rd generation | 40-49 | 1192 | 5377 | 22.17 |
| 2009 | COC 4th generation | 40-49 | 886 | 5377 | 16.48 |
| 2009 | IUD with progestogen | 40-49 | 1952 | 5377 | 36.30 |
| 2009 | Progestin-only pill | 40-49 | 157 | 5377 | 2.92 |
| 2010 | Anti-androgenic COC | 40-49 | 125 | 5725 | 2.18 |
| 2010 | CHC patch | 40-49 | 284 | 5725 | 4.96 |
| 2010 | CHC ring | 40-49 | 472 | 5725 | 8.24 |
| 2010 | COC 2nd generation | 40-49 | 202 | 5725 | 3.53 |
| 2010 | COC 3rd generation | 40-49 | 1141 | 5725 | 19.93 |
| 2010 | COC 4th generation | 40-49 | 939 | 5725 | 16.40 |
| 2010 | Estradiol COC | 40-49 | 25 | 5725 | 0.44 |
| 2010 | IUD with progestogen | 40-49 | 2349 | 5725 | 41.03 |
| 2010 | Progestin-only pill | 40-49 | 188 | 5725 | 3.28 |
| 2011 | Anti-androgenic COC | 40-49 | 106 | 6125 | 1.73 |
| 2011 | CHC patch | 40-49 | 301 | 6125 | 4.91 |
| 2011 | CHC ring | 40-49 | 516 | 6125 | 8.42 |
| 2011 | COC 2nd generation | 40-49 | 184 | 6125 | 3.00 |
| 2011 | COC 3rd generation | 40-49 | 1122 | 6125 | 18.32 |
| 2011 | COC 4th generation | 40-49 | 1001 | 6125 | 16.34 |
| 2011 | Estradiol COC | 40-49 | 43 | 6125 | 0.70 |
| 2011 | IUD with progestogen | 40-49 | 2618 | 6125 | 42.74 |
| 2011 | Progestin-only pill | 40-49 | 234 | 6125 | 3.82 |
| 2012 | Anti-androgenic COC | 40-49 | 115 | 6514 | 1.77 |
| 2012 | CHC patch | 40-49 | 302 | 6514 | 4.64 |
| 2012 | CHC ring | 40-49 | 527 | 6514 | 8.09 |
| 2012 | COC 2nd generation | 40-49 | 164 | 6514 | 2.52 |
| 2012 | COC 3rd generation | 40-49 | 1109 | 6514 | 17.02 |
| 2012 | COC 4th generation | 40-49 | 1121 | 6514 | 17.21 |
| 2012 | Estradiol COC | 40-49 | 68 | 6514 | 1.04 |
| 2012 | IUD with progestogen | 40-49 | 2878 | 6514 | 44.18 |
| 2012 | Progestin-only pill | 40-49 | 230 | 6514 | 3.53 |
| 2013 | Anti-androgenic COC | 40-49 | 110 | 6956 | 1.58 |
| 2013 | CHC patch | 40-49 | 261 | 6956 | 3.75 |
| 2013 | CHC ring | 40-49 | 553 | 6956 | 7.95 |
| 2013 | COC 2nd generation | 40-49 | 150 | 6956 | 2.16 |
| 2013 | COC 3rd generation | 40-49 | 1067 | 6956 | 15.34 |
| 2013 | COC 4th generation | 40-49 | 1095 | 6956 | 15.74 |
| 2013 | Estradiol COC | 40-49 | 73 | 6956 | 1.05 |
| 2013 | IUD with progestogen | 40-49 | 3394 | 6956 | 48.79 |
| 2013 | Progestin-only pill | 40-49 | 253 | 6956 | 3.64 |
| 2014 | Anti-androgenic COC | 40-49 | 103 | 7172 | 1.44 |
| 2014 | CHC patch | 40-49 | 243 | 7172 | 3.39 |
| 2014 | CHC ring | 40-49 | 519 | 7172 | 7.24 |
| 2014 | COC 2nd generation | 40-49 | 153 | 7172 | 2.13 |
| 2014 | COC 3rd generation | 40-49 | 997 | 7172 | 13.90 |
| 2014 | COC 4th generation | 40-49 | 971 | 7172 | 13.54 |
| 2014 | Estradiol COC | 40-49 | 66 | 7172 | 0.92 |
| 2014 | IUD with progestogen | 40-49 | 3858 | 7172 | 53.79 |
| 2014 | Implant | 40-49 | 4 | 7172 | 0.06 |
| 2014 | Progestin-only pill | 40-49 | 258 | 7172 | 3.60 |
| 2015 | Anti-androgenic COC | 40-49 | 100 | 7575 | 1.32 |
| 2015 | CHC patch | 40-49 | 232 | 7575 | 3.06 |
| 2015 | CHC ring | 40-49 | 499 | 7575 | 6.59 |
| 2015 | COC 2nd generation | 40-49 | 136 | 7575 | 1.80 |
| 2015 | COC 3rd generation | 40-49 | 879 | 7575 | 11.60 |
| 2015 | COC 4th generation | 40-49 | 952 | 7575 | 12.57 |
| 2015 | Estradiol COC | 40-49 | 68 | 7575 | 0.90 |
| 2015 | IUD with progestogen | 40-49 | 4409 | 7575 | 58.20 |
| 2015 | Implant | 40-49 | 30 | 7575 | 0.40 |
| 2015 | Progestin-only pill | 40-49 | 270 | 7575 | 3.56 |
| 2016 | Anti-androgenic COC | 40-49 | 76 | 8114 | 0.94 |
| 2016 | CHC patch | 40-49 | 256 | 8114 | 3.16 |
| 2016 | CHC ring | 40-49 | 470 | 8114 | 5.79 |
| 2016 | COC 2nd generation | 40-49 | 116 | 8114 | 1.43 |
| 2016 | COC 3rd generation | 40-49 | 866 | 8114 | 10.67 |
| 2016 | COC 4th generation | 40-49 | 925 | 8114 | 11.40 |
| 2016 | Estradiol COC | 40-49 | 62 | 8114 | 0.76 |
| 2016 | IUD with progestogen | 40-49 | 4976 | 8114 | 61.33 |
| 2016 | Implant | 40-49 | 58 | 8114 | 0.71 |
| 2016 | Progestin-only pill | 40-49 | 309 | 8114 | 3.81 |
| 2017 | Anti-androgenic COC | 40-49 | 75 | 8651 | 0.87 |
| 2017 | CHC patch | 40-49 | 240 | 8651 | 2.77 |
| 2017 | CHC ring | 40-49 | 488 | 8651 | 5.64 |
| 2017 | COC 2nd generation | 40-49 | 129 | 8651 | 1.49 |
| 2017 | COC 3rd generation | 40-49 | 858 | 8651 | 9.92 |
| 2017 | COC 4th generation | 40-49 | 952 | 8651 | 11.00 |
| 2017 | Estradiol COC | 40-49 | 55 | 8651 | 0.64 |
| 2017 | IUD with progestogen | 40-49 | 5442 | 8651 | 62.91 |
| 2017 | Implant | 40-49 | 72 | 8651 | 0.83 |
| 2017 | Progestin-only pill | 40-49 | 340 | 8651 | 3.93 |
| 2018 | Anti-androgenic COC | 40-49 | 64 | 9096 | 0.70 |
| 2018 | CHC patch | 40-49 | 217 | 9096 | 2.39 |
| 2018 | CHC ring | 40-49 | 479 | 9096 | 5.27 |
| 2018 | COC 2nd generation | 40-49 | 121 | 9096 | 1.33 |
| 2018 | COC 3rd generation | 40-49 | 893 | 9096 | 9.82 |
| 2018 | COC 4th generation | 40-49 | 977 | 9096 | 10.74 |
| 2018 | Estradiol COC | 40-49 | 45 | 9096 | 0.49 |
| 2018 | IUD with progestogen | 40-49 | 5811 | 9096 | 63.89 |
| 2018 | Implant | 40-49 | 95 | 9096 | 1.04 |
| 2018 | Progestin-only pill | 40-49 | 394 | 9096 | 4.33 |
| 2019 | Anti-androgenic COC | 40-49 | 65 | 9452 | 0.69 |
| 2019 | CHC patch | 40-49 | 214 | 9452 | 2.26 |
| 2019 | CHC ring | 40-49 | 492 | 9452 | 5.21 |
| 2019 | COC 2nd generation | 40-49 | 99 | 9452 | 1.05 |
| 2019 | COC 3rd generation | 40-49 | 855 | 9452 | 9.05 |
| 2019 | COC 4th generation | 40-49 | 997 | 9452 | 10.55 |
| 2019 | Estradiol COC | 40-49 | 46 | 9452 | 0.49 |
| 2019 | IUD with progestogen | 40-49 | 6153 | 9452 | 65.10 |
| 2019 | Implant | 40-49 | 103 | 9452 | 1.09 |
| 2019 | Progestin-only pill | 40-49 | 428 | 9452 | 4.53 |
| 2020 | Anti-androgenic COC | 40-49 | 51 | 9494 | 0.54 |
| 2020 | CHC patch | 40-49 | 211 | 9494 | 2.22 |
| 2020 | CHC ring | 40-49 | 465 | 9494 | 4.90 |
| 2020 | COC 2nd generation | 40-49 | 46 | 9494 | 0.48 |
| 2020 | COC 3rd generation | 40-49 | 837 | 9494 | 8.82 |
| 2020 | COC 4th generation | 40-49 | 938 | 9494 | 9.88 |
| 2020 | Estradiol COC | 40-49 | 51 | 9494 | 0.54 |
| 2020 | IUD with progestogen | 40-49 | 6299 | 9494 | 66.35 |
| 2020 | Implant | 40-49 | 112 | 9494 | 1.18 |
| 2020 | Progestin-only pill | 40-49 | 484 | 9494 | 5.10 |
| 2021 | Anti-androgenic COC | 40-49 | 46 | 9375 | 0.49 |
| 2021 | CHC patch | 40-49 | 195 | 9375 | 2.08 |
| 2021 | CHC ring | 40-49 | 430 | 9375 | 4.59 |
| 2021 | COC 2nd generation | 40-49 | 38 | 9375 | 0.41 |
| 2021 | COC 3rd generation | 40-49 | 778 | 9375 | 8.30 |
| 2021 | COC 4th generation | 40-49 | 829 | 9375 | 8.84 |
| 2021 | Estradiol COC | 40-49 | 40 | 9375 | 0.43 |
| 2021 | IUD with progestogen | 40-49 | 6403 | 9375 | 68.30 |
| 2021 | Implant | 40-49 | 122 | 9375 | 1.30 |
| 2021 | Progestin-only pill | 40-49 | 494 | 9375 | 5.27 |
| 2022 | Anti-androgenic COC | 40-49 | 39 | 9489 | 0.41 |
| 2022 | CHC patch | 40-49 | 165 | 9489 | 1.74 |
| 2022 | CHC ring | 40-49 | 415 | 9489 | 4.37 |
| 2022 | COC 2nd generation | 40-49 | 27 | 9489 | 0.28 |
| 2022 | COC 3rd generation | 40-49 | 723 | 9489 | 7.62 |
| 2022 | COC 4th generation | 40-49 | 820 | 9489 | 8.64 |
| 2022 | Estradiol COC | 40-49 | 66 | 9489 | 0.70 |
| 2022 | IUD with progestogen | 40-49 | 6558 | 9489 | 69.11 |
| 2022 | Implant | 40-49 | 113 | 9489 | 1.19 |
| 2022 | Progestin-only pill | 40-49 | 563 | 9489 | 5.93 |
| 2004 | Anti-androgenic COC | 50-55 | 5 | 107 | 4.67 |
| 2004 | CHC patch | 50-55 | 6 | 107 | 5.61 |
| 2004 | COC 2nd generation | 50-55 | 16 | 107 | 14.95 |
| 2004 | COC 3rd generation | 50-55 | 68 | 107 | 63.55 |
| 2004 | COC 4th generation | 50-55 | 9 | 107 | 8.41 |
| 2004 | Progestin-only pill | 50-55 | 3 | 107 | 2.80 |
| 2005 | Anti-androgenic COC | 50-55 | 11 | 185 | 5.95 |
| 2005 | CHC patch | 50-55 | 14 | 185 | 7.57 |
| 2005 | CHC ring | 50-55 | 6 | 185 | 3.24 |
| 2005 | COC 2nd generation | 50-55 | 26 | 185 | 14.05 |
| 2005 | COC 3rd generation | 50-55 | 99 | 185 | 53.51 |
| 2005 | COC 4th generation | 50-55 | 24 | 185 | 12.97 |
| 2005 | IUD with progestogen | 50-55 | 2 | 185 | 1.08 |
| 2005 | Progestin-only pill | 50-55 | 3 | 185 | 1.62 |
| 2006 | Anti-androgenic COC | 50-55 | 11 | 217 | 5.07 |
| 2006 | CHC patch | 50-55 | 16 | 217 | 7.37 |
| 2006 | CHC ring | 50-55 | 7 | 217 | 3.23 |
| 2006 | COC 2nd generation | 50-55 | 30 | 217 | 13.82 |
| 2006 | COC 3rd generation | 50-55 | 103 | 217 | 47.47 |
| 2006 | COC 4th generation | 50-55 | 28 | 217 | 12.90 |
| 2006 | IUD with progestogen | 50-55 | 19 | 217 | 8.76 |
| 2006 | Progestin-only pill | 50-55 | 3 | 217 | 1.38 |
| 2007 | Anti-androgenic COC | 50-55 | 8 | 248 | 3.23 |
| 2007 | CHC patch | 50-55 | 15 | 248 | 6.05 |
| 2007 | CHC ring | 50-55 | 6 | 248 | 2.42 |
| 2007 | COC 2nd generation | 50-55 | 35 | 248 | 14.11 |
| 2007 | COC 3rd generation | 50-55 | 109 | 248 | 43.95 |
| 2007 | COC 4th generation | 50-55 | 35 | 248 | 14.11 |
| 2007 | IUD with progestogen | 50-55 | 37 | 248 | 14.92 |
| 2007 | Progestin-only pill | 50-55 | 3 | 248 | 1.21 |
| 2008 | Anti-androgenic COC | 50-55 | 11 | 317 | 3.47 |
| 2008 | CHC patch | 50-55 | 20 | 317 | 6.31 |
| 2008 | CHC ring | 50-55 | 16 | 317 | 5.05 |
| 2008 | COC 2nd generation | 50-55 | 36 | 317 | 11.36 |
| 2008 | COC 3rd generation | 50-55 | 97 | 317 | 30.60 |
| 2008 | COC 4th generation | 50-55 | 39 | 317 | 12.30 |
| 2008 | IUD with progestogen | 50-55 | 88 | 317 | 27.76 |
| 2008 | Progestin-only pill | 50-55 | 10 | 317 | 3.15 |
| 2009 | Anti-androgenic COC | 50-55 | 9 | 433 | 2.08 |
| 2009 | CHC patch | 50-55 | 13 | 433 | 3.00 |
| 2009 | CHC ring | 50-55 | 22 | 433 | 5.08 |
| 2009 | COC 2nd generation | 50-55 | 36 | 433 | 8.31 |
| 2009 | COC 3rd generation | 50-55 | 124 | 433 | 28.64 |
| 2009 | COC 4th generation | 50-55 | 49 | 433 | 11.32 |
| 2009 | IUD with progestogen | 50-55 | 172 | 433 | 39.72 |
| 2009 | Progestin-only pill | 50-55 | 8 | 433 | 1.85 |
| 2010 | Anti-androgenic COC | 50-55 | 11 | 596 | 1.85 |
| 2010 | CHC patch | 50-55 | 16 | 596 | 2.68 |
| 2010 | CHC ring | 50-55 | 28 | 596 | 4.70 |
| 2010 | COC 2nd generation | 50-55 | 40 | 596 | 6.71 |
| 2010 | COC 3rd generation | 50-55 | 144 | 596 | 24.16 |
| 2010 | COC 4th generation | 50-55 | 55 | 596 | 9.23 |
| 2010 | Estradiol COC | 50-55 | 7 | 596 | 1.17 |
| 2010 | IUD with progestogen | 50-55 | 286 | 596 | 47.99 |
| 2010 | Progestin-only pill | 50-55 | 9 | 596 | 1.51 |
| 2011 | Anti-androgenic COC | 50-55 | 12 | 735 | 1.63 |
| 2011 | CHC patch | 50-55 | 19 | 735 | 2.59 |
| 2011 | CHC ring | 50-55 | 32 | 735 | 4.35 |
| 2011 | COC 2nd generation | 50-55 | 39 | 735 | 5.31 |
| 2011 | COC 3rd generation | 50-55 | 134 | 735 | 18.23 |
| 2011 | COC 4th generation | 50-55 | 72 | 735 | 9.80 |
| 2011 | Estradiol COC | 50-55 | 1 | 735 | 0.14 |
| 2011 | IUD with progestogen | 50-55 | 414 | 735 | 56.33 |
| 2011 | Progestin-only pill | 50-55 | 12 | 735 | 1.63 |
| 2012 | Anti-androgenic COC | 50-55 | 7 | 842 | 0.83 |
| 2012 | CHC patch | 50-55 | 14 | 842 | 1.66 |
| 2012 | CHC ring | 50-55 | 28 | 842 | 3.33 |
| 2012 | COC 2nd generation | 50-55 | 34 | 842 | 4.04 |
| 2012 | COC 3rd generation | 50-55 | 138 | 842 | 16.39 |
| 2012 | COC 4th generation | 50-55 | 73 | 842 | 8.67 |
| 2012 | Estradiol COC | 50-55 | 12 | 842 | 1.43 |
| 2012 | IUD with progestogen | 50-55 | 515 | 842 | 61.16 |
| 2012 | Progestin-only pill | 50-55 | 21 | 842 | 2.49 |
| 2013 | Anti-androgenic COC | 50-55 | 7 | 955 | 0.73 |
| 2013 | CHC patch | 50-55 | 20 | 955 | 2.09 |
| 2013 | CHC ring | 50-55 | 22 | 955 | 2.30 |
| 2013 | COC 2nd generation | 50-55 | 32 | 955 | 3.35 |
| 2013 | COC 3rd generation | 50-55 | 135 | 955 | 14.14 |
| 2013 | COC 4th generation | 50-55 | 73 | 955 | 7.64 |
| 2013 | Estradiol COC | 50-55 | 11 | 955 | 1.15 |
| 2013 | IUD with progestogen | 50-55 | 635 | 955 | 66.49 |
| 2013 | Progestin-only pill | 50-55 | 20 | 955 | 2.09 |
| 2014 | Anti-androgenic COC | 50-55 | 7 | 1080 | 0.65 |
| 2014 | CHC patch | 50-55 | 16 | 1080 | 1.48 |
| 2014 | CHC ring | 50-55 | 25 | 1080 | 2.31 |
| 2014 | COC 2nd generation | 50-55 | 23 | 1080 | 2.13 |
| 2014 | COC 3rd generation | 50-55 | 122 | 1080 | 11.30 |
| 2014 | COC 4th generation | 50-55 | 82 | 1080 | 7.59 |
| 2014 | Estradiol COC | 50-55 | 10 | 1080 | 0.93 |
| 2014 | IUD with progestogen | 50-55 | 768 | 1080 | 71.11 |
| 2014 | Progestin-only pill | 50-55 | 27 | 1080 | 2.50 |
| 2015 | Anti-androgenic COC | 50-55 | 5 | 1215 | 0.41 |
| 2015 | CHC patch | 50-55 | 16 | 1215 | 1.32 |
| 2015 | CHC ring | 50-55 | 32 | 1215 | 2.63 |
| 2015 | COC 2nd generation | 50-55 | 28 | 1215 | 2.30 |
| 2015 | COC 3rd generation | 50-55 | 119 | 1215 | 9.79 |
| 2015 | COC 4th generation | 50-55 | 83 | 1215 | 6.83 |
| 2015 | Estradiol COC | 50-55 | 3 | 1215 | 0.25 |
| 2015 | IUD with progestogen | 50-55 | 898 | 1215 | 73.91 |
| 2015 | Progestin-only pill | 50-55 | 31 | 1215 | 2.55 |
| 2016 | Anti-androgenic COC | 50-55 | 7 | 1342 | 0.52 |
| 2016 | CHC patch | 50-55 | 19 | 1342 | 1.42 |
| 2016 | CHC ring | 50-55 | 35 | 1342 | 2.61 |
| 2016 | COC 2nd generation | 50-55 | 24 | 1342 | 1.79 |
| 2016 | COC 3rd generation | 50-55 | 85 | 1342 | 6.33 |
| 2016 | COC 4th generation | 50-55 | 73 | 1342 | 5.44 |
| 2016 | Estradiol COC | 50-55 | 6 | 1342 | 0.45 |
| 2016 | IUD with progestogen | 50-55 | 1062 | 1342 | 79.14 |
| 2016 | Implant | 50-55 | 1 | 1342 | 0.07 |
| 2016 | Progestin-only pill | 50-55 | 30 | 1342 | 2.24 |
| 2017 | Anti-androgenic COC | 50-55 | 7 | 1631 | 0.43 |
| 2017 | CHC patch | 50-55 | 22 | 1631 | 1.35 |
| 2017 | CHC ring | 50-55 | 40 | 1631 | 2.45 |
| 2017 | COC 2nd generation | 50-55 | 26 | 1631 | 1.59 |
| 2017 | COC 3rd generation | 50-55 | 78 | 1631 | 4.78 |
| 2017 | COC 4th generation | 50-55 | 79 | 1631 | 4.84 |
| 2017 | Estradiol COC | 50-55 | 9 | 1631 | 0.55 |
| 2017 | IUD with progestogen | 50-55 | 1329 | 1631 | 81.48 |
| 2017 | Implant | 50-55 | 4 | 1631 | 0.25 |
| 2017 | Progestin-only pill | 50-55 | 37 | 1631 | 2.27 |
| 2018 | Anti-androgenic COC | 50-55 | 10 | 1934 | 0.52 |
| 2018 | CHC patch | 50-55 | 27 | 1934 | 1.40 |
| 2018 | CHC ring | 50-55 | 32 | 1934 | 1.65 |
| 2018 | COC 2nd generation | 50-55 | 27 | 1934 | 1.40 |
| 2018 | COC 3rd generation | 50-55 | 70 | 1934 | 3.62 |
| 2018 | COC 4th generation | 50-55 | 86 | 1934 | 4.45 |
| 2018 | Estradiol COC | 50-55 | 7 | 1934 | 0.36 |
| 2018 | IUD with progestogen | 50-55 | 1631 | 1934 | 84.33 |
| 2018 | Implant | 50-55 | 4 | 1934 | 0.21 |
| 2018 | Progestin-only pill | 50-55 | 40 | 1934 | 2.07 |
| 2019 | Anti-androgenic COC | 50-55 | 8 | 2351 | 0.34 |
| 2019 | CHC patch | 50-55 | 29 | 2351 | 1.23 |
| 2019 | CHC ring | 50-55 | 44 | 2351 | 1.87 |
| 2019 | COC 2nd generation | 50-55 | 24 | 2351 | 1.02 |
| 2019 | COC 3rd generation | 50-55 | 92 | 2351 | 3.91 |
| 2019 | COC 4th generation | 50-55 | 101 | 2351 | 4.30 |
| 2019 | Estradiol COC | 50-55 | 6 | 2351 | 0.26 |
| 2019 | IUD with progestogen | 50-55 | 1985 | 2351 | 84.43 |
| 2019 | Implant | 50-55 | 7 | 2351 | 0.30 |
| 2019 | Progestin-only pill | 50-55 | 55 | 2351 | 2.34 |
| 2020 | Anti-androgenic COC | 50-55 | 6 | 2595 | 0.23 |
| 2020 | CHC patch | 50-55 | 29 | 2595 | 1.12 |
| 2020 | CHC ring | 50-55 | 48 | 2595 | 1.85 |
| 2020 | COC 2nd generation | 50-55 | 8 | 2595 | 0.31 |
| 2020 | COC 3rd generation | 50-55 | 98 | 2595 | 3.78 |
| 2020 | COC 4th generation | 50-55 | 102 | 2595 | 3.93 |
| 2020 | Estradiol COC | 50-55 | 8 | 2595 | 0.31 |
| 2020 | IUD with progestogen | 50-55 | 2232 | 2595 | 86.01 |
| 2020 | Implant | 50-55 | 7 | 2595 | 0.27 |
| 2020 | Progestin-only pill | 50-55 | 57 | 2595 | 2.20 |
| 2021 | Anti-androgenic COC | 50-55 | 4 | 2843 | 0.14 |
| 2021 | CHC patch | 50-55 | 25 | 2843 | 0.88 |
| 2021 | CHC ring | 50-55 | 49 | 2843 | 1.72 |
| 2021 | COC 2nd generation | 50-55 | 5 | 2843 | 0.18 |
| 2021 | COC 3rd generation | 50-55 | 105 | 2843 | 3.69 |
| 2021 | COC 4th generation | 50-55 | 99 | 2843 | 3.48 |
| 2021 | Estradiol COC | 50-55 | 12 | 2843 | 0.42 |
| 2021 | IUD with progestogen | 50-55 | 2471 | 2843 | 86.92 |
| 2021 | Implant | 50-55 | 11 | 2843 | 0.39 |
| 2021 | Progestin-only pill | 50-55 | 62 | 2843 | 2.18 |
| 2022 | Anti-androgenic COC | 50-55 | 4 | 3158 | 0.13 |
| 2022 | CHC patch | 50-55 | 24 | 3158 | 0.76 |
| 2022 | CHC ring | 50-55 | 51 | 3158 | 1.61 |
| 2022 | COC 2nd generation | 50-55 | 7 | 3158 | 0.22 |
| 2022 | COC 3rd generation | 50-55 | 93 | 3158 | 2.94 |
| 2022 | COC 4th generation | 50-55 | 111 | 3158 | 3.51 |
| 2022 | Estradiol COC | 50-55 | 17 | 3158 | 0.54 |
| 2022 | IUD with progestogen | 50-55 | 2742 | 3158 | 86.83 |
| 2022 | Implant | 50-55 | 9 | 3158 | 0.28 |
| 2022 | Progestin-only pill | 50-55 | 100 | 3158 | 3.17 |

**Supplementary Table 4.** Characteristics of hormonal contraceptive (HC) user types of interest

|  | **Non-Switchers (n = 17,453)** | **Rapid Switchers (n = 12,929)** | **Rapid Discontinuers (n = 5,362)** |
| --- | --- | --- | --- |
| **Age Group** |  |  |  |
| 15-19 | 2070 (11.71%) | 1719 (9.35%) | 290 (5.41%) |
| 20-29 | 5928 (33.84%) | 9022 (49.07%) | 1401 (26.13%) |
| 30-39 | 4258 (24.09%) | 5586 (30.38%) | 1726 (32.19%) |
| 40-49 | 4895 (27.69%) | 1986 (10.80%) | 1711 (31.91%) |
| 50-55 | 472 (2.67%) | 73 (0.40%) | 234 (4.36%) |
| **BMI Category** |  |  |  |
| Underweight | 719 (4.07%) | 906 (4.93%) | 198 (3.69%) |
| Healthy | 10442 (59.07%) | 12026 (65.41%) | 234 (52.85%) |
| Overweight | 4206 (23.79%) | 3605 (19.61%) | 1405 (26.20%) |
| Obese | 2310 (13.07%) | 1849 (10.06%) | 925 (17.25%) |
| **HC Type** |  | *(referring to the pre-switch HC formulation)* |  |
| IUD | 5436 (30.75%) | 691 (3.76%) | 34 (0.63%) |
| POP | 508 (2.87%) | 2934 (15.96%) | 588 (10.97%) |
| Anti-androgenic COC | 425 (2.40%) | 1077 (5.86%) | 221 (4.12%) |
| 2nd gen COC | 441 (2.49%) | 606 (3.30%) | 161 (3.00%) |
| 3rd gen COC | 4861 (27.50%) | 4248 (23.10%) | 1923 (35.86%) |
| 4th gen COC | 4205 (23.79%) | 4138 (22.51%) | 1244 (23.20%) |
| Estradiol COC | 22 (0.12%) | 94 (0.51%) | 24 (0.45%) |
| CHC ring | 845 (4.78%) | 2036 (11.07%) | 539 (10.05%) |
| CHC patch | 805 (4.55%) | 2519 (13.70%) | 628 (11.71%) |
| Implant | 129 (0. 73%) | 43 (0.23%) | 0 (0.00%) |

**Supplementary Table 5.** Total number of rapid switchers with diagnosis from curated list of diagnoses related to potential side effects between two hormonal contraceptive (HC) formulations

| **HC_type** | **ICD_10_Code_main** | **count** | **total_count** |
| --- | --- | --- | --- |
| Anti-androgenic COC | B37 | 28 | 323 |
| Anti-androgenic COC | N76 | 25 | 350 |
| Anti-androgenic COC | N92 | 23 | 556 |
| Anti-androgenic COC | L70 | 22 | 180 |
| Anti-androgenic COC | N30 | 21 | 327 |
| Anti-androgenic COC | E28 | 19 | 257 |
| Anti-androgenic COC | N72 | 16 | 176 |
| Anti-androgenic COC | R10 | 12 | 231 |
| Anti-androgenic COC | N91 | 10 | 138 |
| Anti-androgenic COC | F32 | 9 | 140 |
| Anti-androgenic COC | F41 | 8 | 93 |
| Anti-androgenic COC | N94 | 8 | 147 |
| Anti-androgenic COC | G44 | 6 | 81 |
| Anti-androgenic COC | N87 | 6 | 151 |
| Anti-androgenic COC | G43 | 5 | 98 |
| Anti-androgenic COC | N83 | 5 | 127 |
| Anti-androgenic COC | D27 | 4 | 49 |
| Anti-androgenic COC | N64 | 4 | 28 |
| Anti-androgenic COC | N84 | 4 | 64 |
| Anti-androgenic COC | R51 | 4 | 74 |
| Anti-androgenic COC | K80 | 3 | 23 |
| Anti-androgenic COC | N93 | 2 | 63 |
| Anti-androgenic COC | D25 | 1 | 78 |
| Anti-androgenic COC | R53 | 1 | 55 |
| CHC patch | N92 | 59 | 556 |
| CHC patch | N30 | 51 | 327 |
| CHC patch | B37 | 48 | 323 |
| CHC patch | N76 | 45 | 350 |
| CHC patch | R10 | 35 | 231 |
| CHC patch | E28 | 27 | 257 |
| CHC patch | N72 | 27 | 176 |
| CHC patch | F32 | 26 | 140 |
| CHC patch | N87 | 25 | 151 |
| CHC patch | N94 | 24 | 147 |
| CHC patch | L70 | 21 | 180 |
| CHC patch | N83 | 19 | 127 |
| CHC patch | F41 | 15 | 93 |
| CHC patch | G43 | 13 | 98 |
| CHC patch | N91 | 11 | 138 |
| CHC patch | R51 | 10 | 74 |
| CHC patch | D25 | 9 | 78 |
| CHC patch | R53 | 6 | 55 |
| CHC patch | D27 | 5 | 49 |
| CHC patch | G44 | 4 | 81 |
| CHC patch | N93 | 4 | 63 |
| CHC patch | N64 | 2 | 28 |
| CHC patch | N84 | 2 | 64 |
| CHC patch | K80 | 1 | 23 |
| CHC patch | N80 | 1 | 40 |
| CHC ring | N92 | 40 | 556 |
| CHC ring | N30 | 39 | 327 |
| CHC ring | N76 | 38 | 350 |
| CHC ring | B37 | 30 | 323 |
| CHC ring | R10 | 27 | 231 |
| CHC ring | E28 | 23 | 257 |
| CHC ring | N87 | 19 | 151 |
| CHC ring | G43 | 15 | 98 |
| CHC ring | L70 | 15 | 180 |
| CHC ring | N72 | 15 | 176 |
| CHC ring | F32 | 13 | 140 |
| CHC ring | N94 | 13 | 147 |
| CHC ring | N91 | 12 | 138 |
| CHC ring | D25 | 11 | 78 |
| CHC ring | F41 | 11 | 93 |
| CHC ring | N83 | 10 | 127 |
| CHC ring | R51 | 10 | 74 |
| CHC ring | G44 | 8 | 81 |
| CHC ring | N93 | 8 | 63 |
| CHC ring | R53 | 6 | 55 |
| CHC ring | N84 | 5 | 64 |
| CHC ring | K80 | 3 | 23 |
| CHC ring | N80 | 3 | 40 |
| CHC ring | D27 | 1 | 49 |
| COC 2nd generation | N92 | 22 | 556 |
| COC 2nd generation | N76 | 19 | 350 |
| COC 2nd generation | E28 | 13 | 257 |
| COC 2nd generation | N94 | 11 | 147 |
| COC 2nd generation | B37 | 10 | 323 |
| COC 2nd generation | L70 | 10 | 180 |
| COC 2nd generation | R10 | 9 | 231 |
| COC 2nd generation | N72 | 8 | 176 |
| COC 2nd generation | N91 | 8 | 138 |
| COC 2nd generation | N30 | 7 | 327 |
| COC 2nd generation | F32 | 6 | 140 |
| COC 2nd generation | F41 | 6 | 93 |
| COC 2nd generation | R51 | 6 | 74 |
| COC 2nd generation | G43 | 5 | 98 |
| COC 2nd generation | N84 | 5 | 64 |
| COC 2nd generation | D25 | 4 | 78 |
| COC 2nd generation | R53 | 4 | 55 |
| COC 2nd generation | G44 | 3 | 81 |
| COC 2nd generation | N83 | 2 | 127 |
| COC 2nd generation | N93 | 2 | 63 |
| COC 2nd generation | N80 | 1 | 40 |
| COC 2nd generation | N87 | 1 | 151 |
| COC 3rd generation | N92 | 158 | 556 |
| COC 3rd generation | N30 | 87 | 327 |
| COC 3rd generation | B37 | 84 | 323 |
| COC 3rd generation | E28 | 81 | 257 |
| COC 3rd generation | N76 | 81 | 350 |
| COC 3rd generation | R10 | 57 | 231 |
| COC 3rd generation | L70 | 46 | 180 |
| COC 3rd generation | N72 | 45 | 176 |
| COC 3rd generation | F32 | 36 | 140 |
| COC 3rd generation | N91 | 33 | 138 |
| COC 3rd generation | N94 | 33 | 147 |
| COC 3rd generation | N87 | 31 | 151 |
| COC 3rd generation | G44 | 27 | 81 |
| COC 3rd generation | G43 | 25 | 98 |
| COC 3rd generation | N83 | 24 | 127 |
| COC 3rd generation | D25 | 20 | 78 |
| COC 3rd generation | R51 | 20 | 74 |
| COC 3rd generation | F41 | 18 | 93 |
| COC 3rd generation | N93 | 16 | 63 |
| COC 3rd generation | N80 | 15 | 40 |
| COC 3rd generation | D27 | 14 | 49 |
| COC 3rd generation | N84 | 14 | 64 |
| COC 3rd generation | R53 | 13 | 55 |
| COC 3rd generation | N64 | 10 | 28 |
| COC 3rd generation | L68 | 3 | 5 |
| COC 3rd generation | F52 | 1 | 2 |
| COC 3rd generation | K80 | 1 | 23 |
| COC 4th generation | N92 | 115 | 556 |
| COC 4th generation | N76 | 82 | 350 |
| COC 4th generation | N30 | 79 | 327 |
| COC 4th generation | B37 | 63 | 323 |
| COC 4th generation | E28 | 55 | 257 |
| COC 4th generation | L70 | 51 | 180 |
| COC 4th generation | R10 | 49 | 231 |
| COC 4th generation | N94 | 44 | 147 |
| COC 4th generation | N72 | 36 | 176 |
| COC 4th generation | N87 | 35 | 151 |
| COC 4th generation | F32 | 29 | 140 |
| COC 4th generation | N91 | 27 | 138 |
| COC 4th generation | N83 | 26 | 127 |
| COC 4th generation | G44 | 21 | 81 |
| COC 4th generation | R53 | 19 | 55 |
| COC 4th generation | F41 | 18 | 93 |
| COC 4th generation | D25 | 17 | 78 |
| COC 4th generation | G43 | 17 | 98 |
| COC 4th generation | D27 | 16 | 49 |
| COC 4th generation | N93 | 14 | 63 |
| COC 4th generation | N84 | 12 | 64 |
| COC 4th generation | N80 | 10 | 40 |
| COC 4th generation | R51 | 10 | 74 |
| COC 4th generation | K80 | 8 | 23 |
| COC 4th generation | N64 | 8 | 28 |
| COC 4th generation | L68 | 1 | 5 |
| Estradiol COC | B37 | 2 | 323 |
| Estradiol COC | D25 | 2 | 78 |
| Estradiol COC | N76 | 2 | 350 |
| Estradiol COC | N92 | 2 | 556 |
| Estradiol COC | F32 | 1 | 140 |
| Estradiol COC | L70 | 1 | 180 |
| Estradiol COC | N30 | 1 | 327 |
| Estradiol COC | N83 | 1 | 127 |
| Estradiol COC | N84 | 1 | 64 |
| Estradiol COC | N87 | 1 | 151 |
| Estradiol COC | N91 | 1 | 138 |
| Estradiol COC | N93 | 1 | 63 |
| Estradiol COC | R10 | 1 | 231 |
| IUD with progestogen | N92 | 24 | 556 |
| IUD with progestogen | R10 | 12 | 231 |
| IUD with progestogen | N83 | 11 | 127 |
| IUD with progestogen | N84 | 7 | 64 |
| IUD with progestogen | D25 | 5 | 78 |
| IUD with progestogen | N72 | 4 | 176 |
| IUD with progestogen | N87 | 4 | 151 |
| IUD with progestogen | B37 | 3 | 323 |
| IUD with progestogen | E28 | 3 | 257 |
| IUD with progestogen | N76 | 3 | 350 |
| IUD with progestogen | N93 | 3 | 63 |
| IUD with progestogen | F32 | 2 | 140 |
| IUD with progestogen | N30 | 2 | 327 |
| IUD with progestogen | N94 | 2 | 147 |
| IUD with progestogen | D27 | 1 | 49 |
| IUD with progestogen | F41 | 1 | 93 |
| IUD with progestogen | K80 | 1 | 23 |
| IUD with progestogen | L70 | 1 | 180 |
| IUD with progestogen | N64 | 1 | 28 |
| IUD with progestogen | N91 | 1 | 138 |
| IUD with progestogen | R51 | 1 | 74 |
| Implant | F32 | 1 | 140 |
| Implant | F41 | 1 | 93 |
| Implant | N30 | 1 | 327 |
| Implant | N83 | 1 | 127 |
| Implant | N91 | 1 | 138 |
| Implant | N92 | 1 | 556 |
| Progestin-only pill | N92 | 112 | 556 |
| Progestin-only pill | B37 | 55 | 323 |
| Progestin-only pill | N76 | 55 | 350 |
| Progestin-only pill | N30 | 39 | 327 |
| Progestin-only pill | E28 | 36 | 257 |
| Progestin-only pill | N91 | 34 | 138 |
| Progestin-only pill | N87 | 29 | 151 |
| Progestin-only pill | R10 | 29 | 231 |
| Progestin-only pill | N83 | 28 | 127 |
| Progestin-only pill | N72 | 25 | 176 |
| Progestin-only pill | G43 | 18 | 98 |
| Progestin-only pill | F32 | 17 | 140 |
| Progestin-only pill | F41 | 15 | 93 |
| Progestin-only pill | N84 | 14 | 64 |
| Progestin-only pill | L70 | 13 | 180 |
| Progestin-only pill | N93 | 13 | 63 |
| Progestin-only pill | R51 | 13 | 74 |
| Progestin-only pill | G44 | 12 | 81 |
| Progestin-only pill | N94 | 12 | 147 |
| Progestin-only pill | N80 | 10 | 40 |
| Progestin-only pill | D25 | 9 | 78 |
| Progestin-only pill | D27 | 8 | 49 |
| Progestin-only pill | K80 | 6 | 23 |
| Progestin-only pill | R53 | 6 | 55 |
| Progestin-only pill | N64 | 3 | 28 |
| Progestin-only pill | F52 | 1 | 2 |
| Progestin-only pill | L68 | 1 | 5 |
